# Expanding CIRdb, a comprehensive catalog of whole-exome sequencing data of Canary Islanders

**DOI:** 10.1101/2025.11.24.25340885

**Authors:** Ana Díaz-de Usera, Luis A. Rubio-Rodríguez, Adrián Muñoz-Barrera, Jose M. Lorenzo-Salazar, Beatriz Guillen-Guio, David Jáspez, Almudena Corrales, Itahisa Marcelino-Rodríguez, María Del Cristo Rodríguez-Pérez, Antonio Cabrera-de León, Rafaela González-Montelongo, Raquel Cruz-Guerrero, Ángel Carracedo, Carlos Flores

## Abstract

Within the intricate European genetic diversity landscape, Canary Islanders exhibit a unique genetic admixture, comprising European (EUR), North African (NAF), and sub-Saharan African (SSA) ancestries. This study aimed to comprehensively characterize the full spectrum of small genetic variation among 920 unrelated donors from this population based on whole-exome sequencing data to further develop CIRdb as the Canary Islanders-specific reference catalog of genetic variation. We combined this with SNP array data and whole-genome sequencing for specific analyses, revealing a total of 387,555 variants, of which 15.1% were previously unreported. Notably, 74.4% of these variants were classified as rare (with frequency <0.5%), including up to 40% of singletons. We also identified and curated a set of 2,068 variants prioritized as putative pathogenic. Intriguingly, the novel pathogenic variants exhibited enrichment in respiratory, cardiovascular, and metabolic disorders. Genetic differentiation patterns clustered separately individuals from the smallest islands, providing fine-grained insights into within-archipelago differentiation. A scan of local genetic ancestry deviations across the genome revealed an EUR ancestry enrichment around the 17q21.31 inversion, widely recognized for positive selection and associated to pleiotropic effects across pulmonary, infectious, and immunological diseases. Our results also evidenced a selective sweep shared by Canary Islanders and the NAF population around Prune Exopolyphosphatase 1 gene, which is associated with body mass index, cardiovascular health, and metabolic traits. Taken together, CIRdb presents a valuable resource of exome-wide genetic variation in a population at the edge of Southwestern European genetic diversity.

## Introduction

Democratization of next-generation sequencing has opened the possibility for international efforts aimed at deeply characterizing genetic variation in different human populations to improve biomedical research and clinical practice (Karczewski et al. 2020; Mulder et al. 2018). The 1000 Genomes Project (1KGP) was among the first efforts to foster the discovery of genetic variation in diverse populations revealing that our genome differs in roughly 5-6 million positions from the human reference genome (1000 Genomes Project Consortium et al. 2015). Many studies have evidenced the biomedical consequences of ignoring the genetic background of populations (Ghouse et al. 2018; Sirugo, Williams, and Tishkoff 2019; Wojcik et al. 2019). These and others have served to establish the grounds to increase the diversity in catalogs of genetic variation to optimally represent population-specific particularities and to support Personalized Medicine strategies across populations worldwide (Kore et al. 2025). Many countries have developed their own genetic variation catalogs or have conducted extensive genetic studies in control individuals (Nagasaki et al. 2015; UK10K Consortium et al. 2015; Jeroncic et al. 2016; Scott et al. 2016; Chheda et al. 2017; Kim et al. 2018; Fattahi et al. 2019; Apol et al. 2022) by combining variant information and population features (Wong et al. 2013; Genome of the Netherlands Consortium 2014; Dopazo et al. 2016). In Spain, a pioneer study of 267 healthy individuals from Galicia and Andalusia highlighted the need to establish local population catalogs specifically in this country for an optimal understanding of genetic variation with clinical relevance (Dopazo et al. 2016).

The Canary Islands archipelago (Spain) is located ∼100 km from the nearest point to the northwest African coast, at the southwestern edge of Europe (**Figure 1**). Originally, the Canary Islands were inhabited by aborigines whose most likely ancestral origin was in the Berber population from North Africa (Hooton 1970; Arauna et al. 2017; Serrano et al. 2023). Notwithstanding, during its conquest by Europeans in the 15th century, the archipelago was subject to important events of admixture and displacement of aboriginals by European populations and imported slaves (Maca-Meyer et al. 2005; Rodríguez-Varela et al. 2017; Fregel et al. 2019). As a result, the current Canary Islanders are a genetically admixed population of three main ancestries: European (EUR), North African (NAF), and sub-Saharan African (SSA). Mitogenomic studies in the current inhabitants have further unraveled fine-grained influences such as from Portuguese and Galicians (García-Olivares et al. 2023). Furthermore, the geographic isolation of this population; the effects of sexual asymmetry, evidenced by the historically progressive decrease in male indigenous lineages (Flores et al. 2003; Fregel et al. 2009); and multiple local adaptation events have finally modeled a particular genetic makeup where, on average, 75-83% has an EUR ancestry component, 17-23% has an NAF ancestry component, and 2% or less has an SSA ancestry component (Pino-Yanes et al. 2011; Botigué et al. 2013). Individually, larger African ancestries were more recently recognized, with up to 38.2% NAF and 9.5% SSA (Díaz-de Usera et al. 2022). However, although the Canary Islands populations (aboriginal, historical, and modern) have been subjected to several analyses during the last two decades, the studies have generally focused on a few genetic markers or small sample sizes.

**Figure 1.**
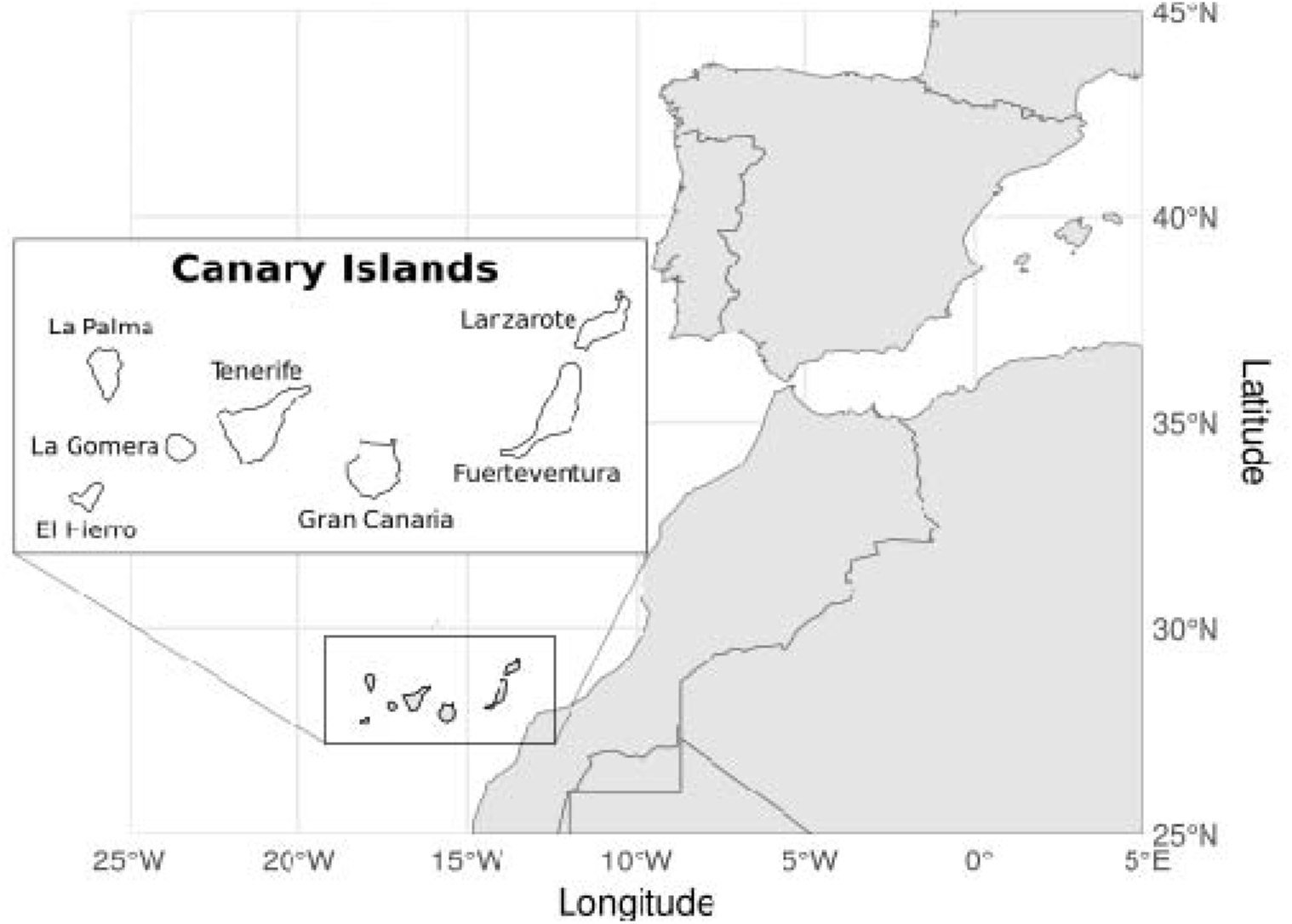
Geographical location of the Canary Islands.

Based on this and the particular genetic makeup of the current Canary Islands populations, we developed the Canary Islanders reference database, termed CIRdb (Díaz-de Usera et al. 2022), as a detailed sequence-based exome reference catalog of genetic variation to allow disentangling population specificities with biomedical impact. In previous studies of the CIRdb cohort, we have exposed the overall design and methods to build the catalog, including population analyses based on SNP arrays and mitogenomes (Díaz-de Usera et al. 2022; García-Olivares et al. 2023). Here, we analyze whole-exome sequencing (WES) data of the cohort to improve the population-specific reference catalog, also involving SNP array data and whole-genome sequencing (WGS) data for specific purposes. This enabled laying the foundation of resources for Genomic Medicine programs in the region and to further deepen into the analysis of the admixture and putative signals of adaptation.

## Materials and Methods

### Sample selection, genotyping, and reference population datasets

The study was approved by the Research Ethics Committee of the Hospital Universitario Nuestra Señora de Candelaria (CHUNSC_2020_95) and performed according to The Code of Ethics of the World Medical Association (Declaration of Helsinki).

The CIRdb cohort was obtained from a total of 1,024 controls that were selected from donors (483 males, 541 females) from the largest general population cohort of the Canary Islands, the ‘CDC of the Canary Islands’ (Cardiovascular, Diabetes, and Cancer) (Cabrera de León et al. 2008). Individuals who self-reported the absence of cardiovascular, metabolic, immunologic, or cancer diseases and had at least two generations of ancestors born on the same island were included in the present study. The island assigned to each individual corresponded to the island of origin of the four grandparents. However, owing to difficulties in sample collection, as was the case for Fuerteventura, this criterion was relaxed, allowing one of the four grandparents to be from a different island in the archipelago. Individuals whose grandparents were not from the same island were classified as No Island Assigned (NIA). These were considered for establishing the catalog, but not for the population, ancestry, or inbreeding analyses conducted in the study. Procedures for DNA extraction, SNP array genotyping, and quality control (QCs) were described previously (Guillen-Guio et al. 2018; Díaz-de Usera et al. 2022). After QCs, 920 samples (439 males, 481 females) with paired WES and SNP array data were kept for further analyses: 106 from El Hierro, 99 from La Palma, 145 from La Gomera, 165 from Tenerife, 215 from Gran Canaria, 47 from Fuerteventura, 114 from Lanzarote, and 29 considered NIA.

Reference WGS data from 1KGP Phase 3 (1000 Genomes Project Consortium et al. 2015) were included in specific analyses to contextualize the genetic composition of the Canary Islanders in the EUR population landscape, which encompassed data from Finnish (FIN) (N =99), British (GBR) (N =91), and Utah Residents with Northern and Western European ancestry (CEU) (N =99). In addition, data from the Iberian population in Spain (IBS) (N =107) was also considered for its genetic proximity to the Canary Islands population. For the African component, the SSA population was selected using the Yoruba population in Ibadan (Nigeria) (YRI) (N =108) in the 1KGP as a proxy. The NAF (N =32) was represented by data from individuals recruited at different locations north of the African continent (Serradell et al. 2024). See the Supplementary Material for further details.

A total of 325,756 variants obtained by WES and 507,607 SNP array variants, a subset of all variants passing the QCs and included in the catalog, were used for the analyses that needed intersection with the population reference datasets. The SNP array data were only used for the ancestry and the Runs of Homozygosity (ROHs) analyses.

### Whole-exome sequencing

Nextera DNA Exome Library Prep with a 350 bp insert size and Illumina DNA Prep with Enrichment kits were used for WES library preparation (Díaz-de Usera et al. 2022; García-Olivares et al. 2023), according to the manufacturer’s instructions (Illumina Inc., San Diego, CA, USA). Pools of up to 12 dual-indexed libraries at 2 nM loading concentration were sequenced on Illumina HiSeq 4000 and NovaSeq 6000 Sequencing Systems (Illumina Inc.) with 75 bp and 100 bp paired-end reads, respectively, following the vendor’s guidance. All experiments used 1% PhiX Control V3 (Illumina Inc.) following the manufacturer’s recommendations.

Genomic data were processed at the TeideHPC Supercomputing facility (https://teidehpc.iter.es/en/home/), following an in-house bioinformatic pipeline described elsewhere (Tosco-Herrera et al. 2022), composed mainly of preprocessing, variant discovery, and QC steps. Additionally, GATK Best Practices (O’Connor and van der Auwera 2020) for large cohorts were implemented in the joint variant calling stage using GATK HaplotypeCaller and the Genomic Variant Call Format (GVCF) mode. After variant calling, different filters were applied to refine the genetic variants included in the catalog (see Supplementary Material).

### Variant annotation

Variant annotation was performed with ANNOVAR v18.04.16 (Wang, Li, and Hakonarson 2010) and The Ensembl Variant Effect Predictor (VEP) v100 (McLaren et al. 2016), using GRCh37/hg19 as the reference genome (Church et al. 2011). The software, databases, and plugins used for variant annotation are listed in Supplementary Table S1.

Pathogenicity was defined according to the InterVar tool and the C-score reported by Combined Annotation Dependent Depletion (CADD) (Rentzsch et al. 2019). The C-score and the mutation significance cutoff (MSC) were used in combination so that the variant was cataloged as putative pathogenic when the C-score was higher than MSC (at 99% confidence). Statistical differences among the number of variants in the different analyses were assessed using the R v4.0.2 environment (R Core Team 2022), based on one-way ANOVA and Duncan’s new multiple range tests (*p*<0.05). In particular cases, a Fisher’s exact test with subsequent Bonferroni correction was also used for variant comparisons. Enrichment analyses were performed using the Genomic Regions Enrichment of Annotations Tool (GREAT) (McLean et al. 2010), Enrichr (Kuleshov et al. 2016), and GeneSCF (Subhash and Kanduri 2016). The annotation of variants and genes with traits and diseases was performed using the GWAS Catalog (Buniello et al. 2019), Open Targets Genetics (Ghoussaini et al. 2021), Open Targets Platform (Ochoa et al. 2021), GeneCards (Safran et al. 2021), and OMIM r20210128 (Amberger et al. 2009).

### Principal Component Analysis and Discriminant Analysis of Principal Components

To adapt to the WES data context, some modifications (i.e., variant exclusions due to call rate <0.80) from the aforementioned QC pipeline were done using PLINK v1.9 (Chang et al. 2015). Principal Component Analysis (PCA) was performed using PLINK v1.9, after excluding variants located in regions of long-range linkage disequilibrium (LD) and with high LD (window size=50, step size=5, r2=0.05), as previously described (Guillen-Guio et al. 2018). For stratified analyses of genetic differentiation based on alternative allele frequency (AAF) range, variants were further sorted by allele frequency in common (AAF>0.05), low frequency (0.005≤ AAF ≤0.05), and rare (AAF<0.005). The same parameters (i.e., sample, variants, and AAF filters) were used for Discriminant Analysis of Principal Components (DAPC) (Jombart, Devillard, and Balloux 2010). Since rare variants consisting of singletons and doubletons behaved as outliers, they were removed from the PCA and DAPC assessments. A nonparametric Mann-Whitney U-test was used to assess the differences between the first three principal components (PCs) adjusting for the number of comparisons.

### Genetic ancestry deviations and runs of homozygosity

Ancestry inferences based on ELAI v1.01 (Guan 2014) were previously obtained for the CIRdb individuals using SNP array data (Díaz-de Usera et al. 2022). Ancestry analyses included WGS data from 423 individuals from the reference populations, which comprised all YRI, NAF, and EUR datasets except the IBS to avoid spurious effects on ancestry estimates due to the genetic proximity between populations (Guillen-Guio et al. 2018; Díaz-de Usera et al. 2022). These estimates were based on a three-way admixture model of EUR, NAF, and SSA, assuming 14 generations since the last admixture event based on our previous observations (Guillen-Guio et al. 2018). Local ancestry block sizes and the average number of ancestry-related blocks in a haploid Canary Islander genome were calculated after removing centromeres and considering the flanking blocks as different elements to avoid overestimation of the block lengths. Deviations in the local parental genomic ancestries were individually assessed for each ancestry using the method developed by Zhu et al. (Zhu 2012). Loci with a Z-score >|3| were considered significantly deviating from the average (Guillen-Guio et al. 2018).

PLINK v1.9-based ROH estimates, obtained according to previous procedures (Kirin et al. 2010; Guillen-Guio et al. 2018), were classified by the average length (0.5-1, 1-2, 2-4, 4-8, 8-16, and ≥16 Mb). We also considered a simplified version of the classification proposed by Pemberton and colleagues (<1.6Mb and ≥1.6Mb) (Pemberton et al. 2012). In addition, the average total length of ROHs and the number of fragments in ROHs were studied per island using nonparametric Mann-Whitney U-tests and Bonferroni correction adjustments (*p*<2.38×10^-3^ to adjust for 21 comparisons).

### Selective sweep analyses around the exonic variant of Prune Exopolyphosphatase 1 (PRUNE1) gene

As part of the DAPC, we first extracted variants contributing more to linear discriminant 1 (LD1) within the common frequency range. To evaluate the likely biomedical implications between ancestry and diseases, the AAF of these prioritized variants was analyzed using Fisher’s exact tests applying a Bonferroni correction to the comparisons between Canary Islanders, EUR, and NAF populations (*p*<3.33×10^-5^ to adjust for 1,501 comparisons). For those variants with statistical differences, the number of individuals was first normalized to match the sample size of the smallest group (i.e., NAF). Subsequently, Weir and Cockerham’s F_ST_ (Weir and Cockerham 1984) and Population Branch Statistics (PBS) (Yi et al. 2010) estimates were obtained for each variant. Next, we used the software iSAFE v1.1.1 (Akbari et al. 2018) to evaluate whether the variant with the highest F_ST_ estimation value in the comparisons with EUR subpopulations and its flanking regions had evidence of selective sweep. An iSAFE scan was run in the region located one megabase (Mb) upstream and downstream from the variant of interest, considering the IgnoreGaps flag and the default MaxFreq value (0.95) (see Supplementary Material). For this analysis, the *Homo sapiens* (GRCh37) ancestral FASTA sequence was obtained from Ensembl release 75 (http://ftp.ensembl.org/pub/release-75/fasta/ancestral_alleles/). Variants with the largest iSAFE scores were prioritized for functional annotation using the Variant to Gene (V2G) score (Ghoussaini et al. 2021) (https://genetics.opentargets.org/).

### Phenotype-Wide Association Study of the PRUNE1 exonic variant

To perform a phenome-wide association study (PheWAS) analysis and given that previous studies support sufficient power for PheWAS of common variants with >200 cases (Verma et al. 2018), we examined associations between the previously identified statistically significant SNP in *PRUNE1* (rs3738476_C) and phenotypes based on the 9^th^ version of the International Classification of Diseases catalog (ICD-9 codes), using the PheWAS v0.99.6.1 R package. All the 920 individuals were mapped to 66 documented diseases in the ICD-9 codes that included at least 20 counts in the cohort and then converted into individual phenotype groups (‘phecodes’). Analyses included age and sex as covariates in all PheWAS models and statistical significance was set at *p*=7.6×10^-4^ to correct for multiple comparisons.

## Results

### Exonic genetic variation in the Canary Islanders

A total of 920 individuals and 387,555 WES variants were finally included in CIRdb. No statistical differences were found in the average number of variants identified per individual across islands (one-way ANOVA test, *p*=0.558) (Supplementary Table S2). Variants were classified into 357,846 single nucleotide variants (SNVs), 18,574 deletions, and 11,135 insertions with an average transition/transversion (Ti/Tv) ratio in autosomes of 2.90 ± 0.04 (SD). The distribution by AAF strata was: 47,493 common, 51,672 low frequency, and 288,390 rare variants. There were 149,851 singletons and 56,188 doubletons in the rare AAF stratum. Thus, 74.4% of the variation had AAF<0.005, and nearly 40% of them was private to individuals. There were statistical differences between islands in the low frequency and rare AAF strata (Supplementary Figure S1) (one-way ANOVA test, *p*<2×10^-16^ for both comparisons; Duncan’s new multiple range tests, *p*<0.05).

Heterozygosity analyses revealed that El Hierro had the highest number of homozygous variants among all the islands (one-way ANOVA test, *p*=1.95×10^-3^). Out of the 120,273 variants found in El Hierro, 43,613 were homozygous for the alternative allele in at least one individual and 2,565 of those variants were found in homozygous state in at least 90% of individuals. We found statistical differences after Bonferroni correction in AAF for 16 variants between El Hierro and some of the other islands (Fisher’s exact test, *p*<1.95×10^-5^) (Supplementary Table S3). All these variants were cataloged as non-pathogenic by the InterVar tool, CADD, and the inferred impact in the Ensembl VEP annotation. Data from Open Targets Genetics linked several of the 16 variants to autoimmune diseases, cardiovascular traits, hair color, and skin browning processes, among others (Supplementary Table S3). One of these variants is the missense variant rs2249265, which is functionally linked to *PTGFRN* and has been associated with type 2 diabetes (T2D) (Vujkovic et al. 2020), showing statistical differences in AAF between El Hierro and La Gomera.

Considering pathogenicity classifications (combining the InterVar tool classification, C-score, and MSC values) and high impact together, 2,068 variants were identified globally as putative pathogenic in CIRdb. These variants were enriched in genes involved in oncological diseases as well as in neurodegenerative and intellectual disability based on Jensen Disease Ontology. A similar pattern was observed in a stratified analysis by island (Supplementary Table S4). Overall, this evidenced enrichment in ciliopathies, neuropathologies, cardiovascular traits, and metabolic disorders. Sensory diseases, mainly related to ocular conditions such as retinitis pigmentosa, cataract and cone-rod or fundus dystrophy, were also significantly enriched (**Figure 2**).

**Figure 2.**
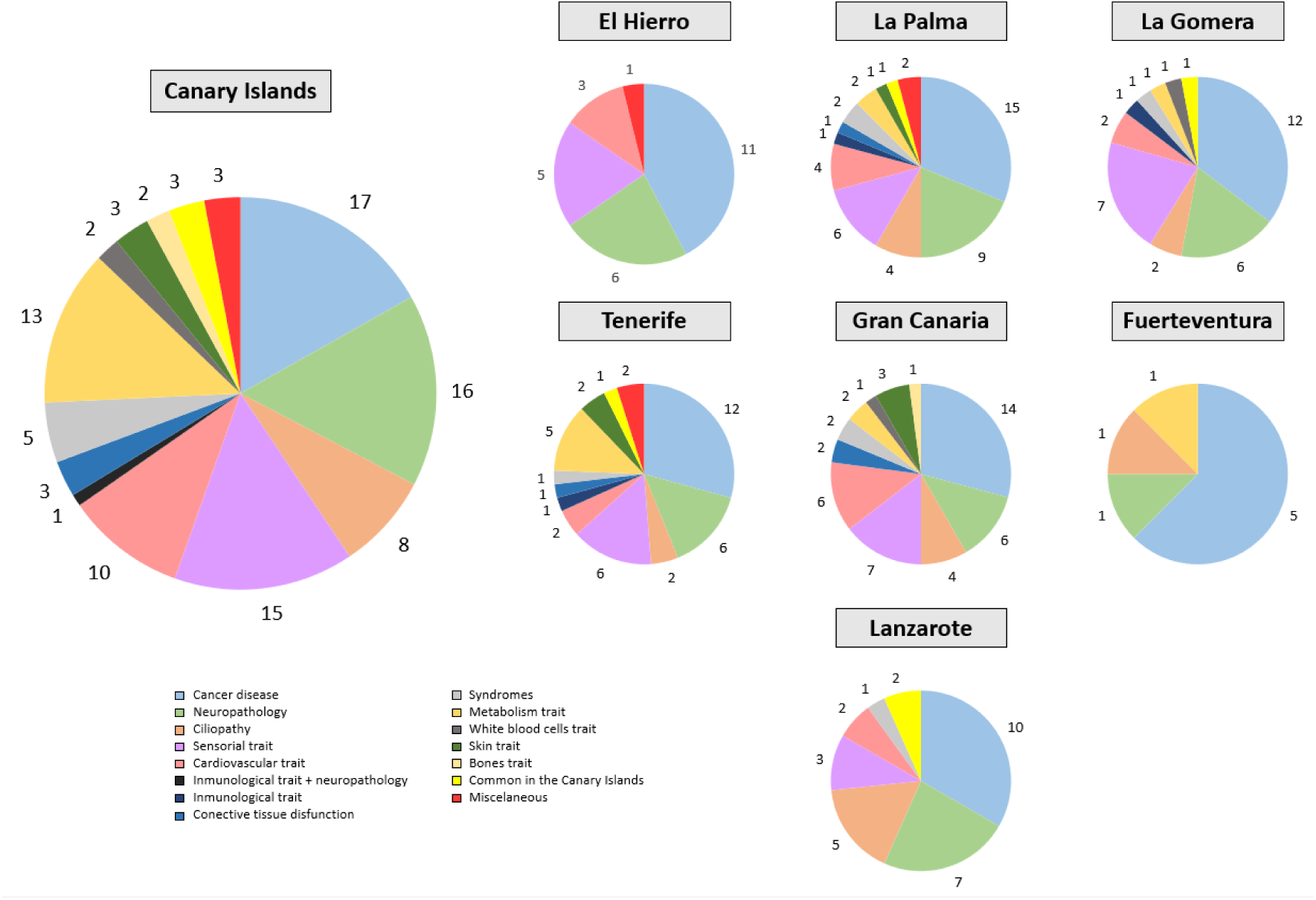
Enrichment analysis on the 2,068 putative pathogenic variants identified in the CIRdb catalog. Annotations in Jensen Disease Ontology were evaluated.

When the affected genomic region and the impact classification provided by Ensembl VEP were simultaneously considered, 74.5% of variants in exonic and intronic regions were within the four different impact categories (77.6% for high impact, 97.4% for moderate, 97.5% for low, and 74.9% for modifier) (**Figure 3**). Splicing with high impact was found for 3,695 variants (17.8% of the total). Moreover, in the splicing regions, nearly one out of every two variants (i.e., 49.1%) were singletons. In line with the idea that variants of potentially higher functional importance are more likely to be singletons (Ke, Taylor, and Cardon 2008), we found a higher proportion of singletons among the high-impact variants (**Figure 4**). Further assessments of missense and loss-of-function variants (LoF) can be found in Supplementary Figure S2 and Supplementary Table S5.

**Figure 3.**
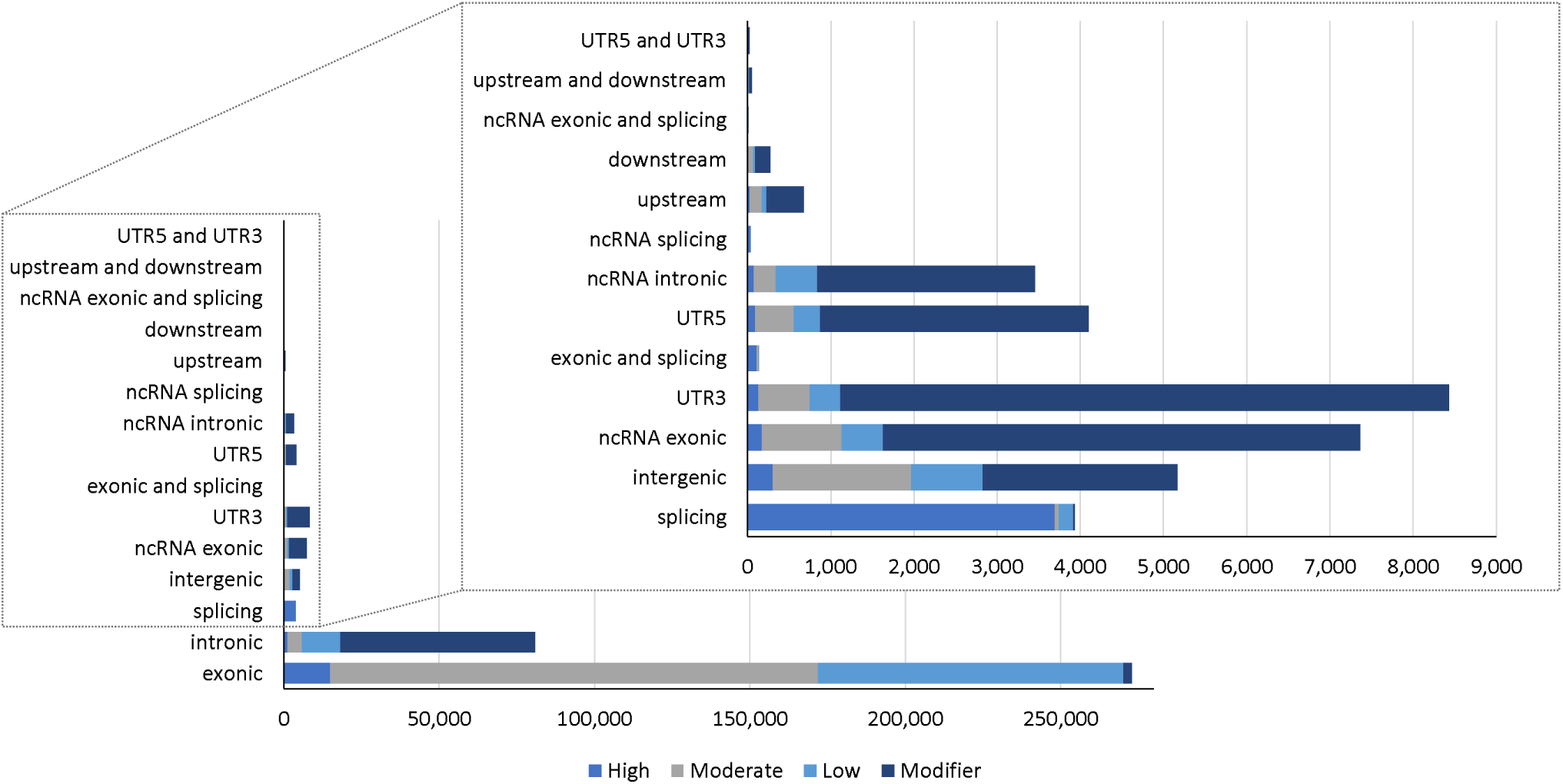
Distribution of whole-exome sequencing variants found in CIRdb based on genomic region and impact classification provided by Ensembl VEP. Colors represent the following impact categories: dark blue for modifier impact, medium blue for high impact, grey for moderate impact, and light blue for low impact. ncRNA, noncoding RNA.

**Figure 4.**
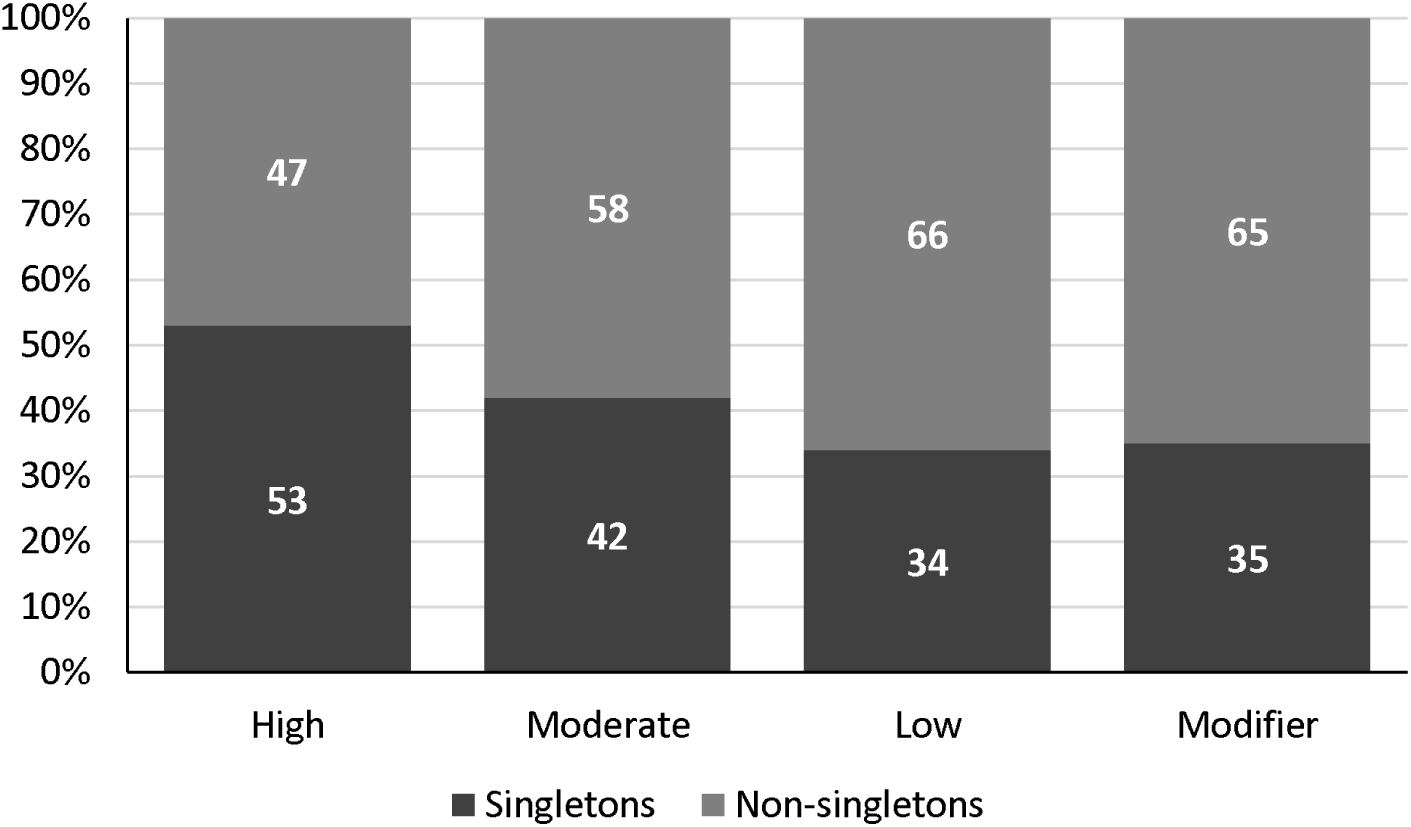
Proportion of singletons and non-singletons across different impact classifications found in CIRdb whole-exome sequencing data.

### Novel variants in the CIRdb catalog

A total of 15.1% of WES variants (n=58,389) were categorized as novel since they were not present in dbSNP154, 1KGP, gnomAD exome v2.1.1 (Karczewski et al. 2020), TOPMed v5 (Taliun et al. 2021), or the HGDP-CEPH (Bergström et al. 2020) projects. La Palma island had the highest average of novel variants per individual, followed by Gran Canaria, Tenerife, La Gomera, Lanzarote, El Hierro, and Fuerteventura. However, statistical differences were observed only when compared with the last two islands (Duncan’s new multiple range test, *p*<0.05). The novel variants were categorized by their AAF strata in 12 common, 1,311 with low frequency, 8,380 rare, 10,019 doubletons, and 38,667 singletons. Thus, 66% of the novel variants were also private to individuals. By impact, novel variants were enriched in the high impact class when compared to all variants identified (**Figure 5**) and were distributed in 19.7% of high impact, 39.7% with moderate, 17.4% with low, and 23.1% classified as modifier. Additionally, 17.5% of the novel variants were also predicted to be LoF, and 1,242 (2.13%) of the novel variants were predicted as putatively pathogenic (Supplementary Table S6). Overall, these variants were in genes associated with cardiovascular, immunological, respiratory, allergic, and sensory diseases, among others, and some of the affected genes were linked to skin pigmentation and sunburn. Furthermore, various novel putatively pathogenic variants affected genes that have been associated with COVID-19 susceptibility and severity, such as *TYK2* and *RAVER1* (COVID-19 Host Genetics Initiative 2023) (Supplementary Table S7).

**Figure 5.**
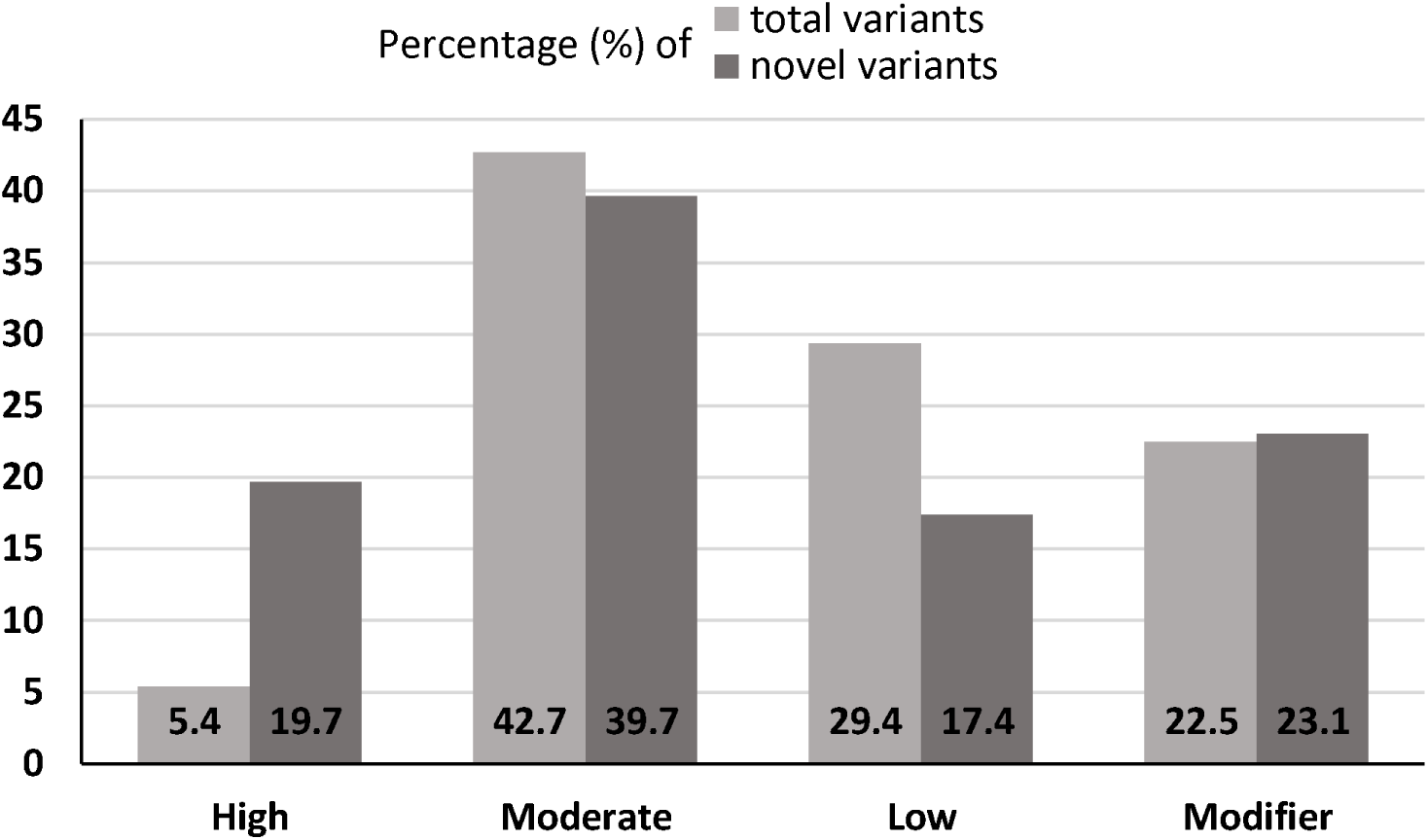
Distribution of total and novel variants based on impact classification found in CIRdb whole-exome sequencing data.

### Shared genetic variation between Canary Islanders and North Africans

We found 10,758 WES variants exclusively shared between the Canary Islanders of CIRdb and the NAF WGS data (i.e., were private to both populations). We found no statistical differences in the number of these variants per island (one-way ANOVA test, *p*=0.456). However, La Palma showed the lowest number of the shared variants (622 ± [SD] 35) in comparison to all the other islands (one-way ANOVA test, *p*=0.002; Duncan’s new multiple range test, *p*<0.05). Fuerteventura, Lanzarote, and El Hierro showed the largest number of these private variants shared with NAF, followed by Tenerife and Gran Canaria. Their AAF classification was: 1,178 common, 1,985 low frequency, 3,442 rare, 1,506 doubletons, and 2,647 singletons. Most of them were classified as of moderate impact (41.3%) and none of these variants were predicted to be putatively pathogenic. Missense, synonymous, and intronic variants were the most abundant categories (Supplementary Table S8). The 50 variants with high impact and CADD>MSC similarly provided benign predictions or were variants of uncertain significance (VUS). Among these 50 variants, 13 showed statistical differences in AAF between the Canary Islands considered altogether and the NAF population after Bonferroni correction (Fisher’s exact test, *p*<0.001) (Supplementary Table S9). The AAF of five of these 13 variants was statistically different between some Canary Islands populations and NAF. Interestingly, we did not find AAF differences for any of the 13 variants between Fuerteventura (one of the closest islands to the northwest African coast) and NAF. Nine out of these 13 variants were exonic to *GPRC6A*, *RP1L1*, *OR4X2*, *PKD1L2*, *KRT38*, *SIRPA*, and *DEFB126*. Interestingly, a genome-wide association study previously linked *DEFB126* variation with coronary artery disease in type 1 diabetes (Antikainen et al. 2021).

### Exome-wide genetic differentiation of the Canary Islands populations

In the common frequency strata (AAF>0.05), the first two main PCs clustered the Canary Islanders into three different groups: the first encompassed individuals from La Gomera, the second included the individuals from El Hierro, and the third encompassed individuals from the other five islands (Mann-Whitney U-test, *p*<2.50×10^-16^ for all comparisons) (**Figure 6A**). In the rare AAF strata (AAF<0.005), the third main PC evidenced the differentiation of individuals from Gran Canaria from the rest of the islands (Mann-Whitney U-test, *p*<2.20×10^-16^ for the comparison of Gran Canaria against the rest of islands) (**Figure 6B**). When comparing against the reference populations, the Canary Islanders showed an intermediate position between the EUR and NAF populations (Mann-Whitney U-test, *p*<1×10^-22^ for all comparisons in PC1) as observed elsewhere (Guillen-Guio et al. 2018; Díaz-de Usera et al. 2022) (Supplementary Table S10). Tenerife and La Palma exhibited greater affinities with the EUR population, eminently with IBS, whereas La Gomera was the island clustering closer to NAF. Nevertheless, all pairwise comparisons among islands and EUR populations were significant (*p*<1×10^-21^). El Hierro, La Gomera, and NAF differentiated from the rest of populations for the first three PCs (Mann-Whitney U-test, *p*<6×10^-6^), except for the comparison of La Gomera and NAF in PC2 (Supplementary Table S10).

**Figure 6.**
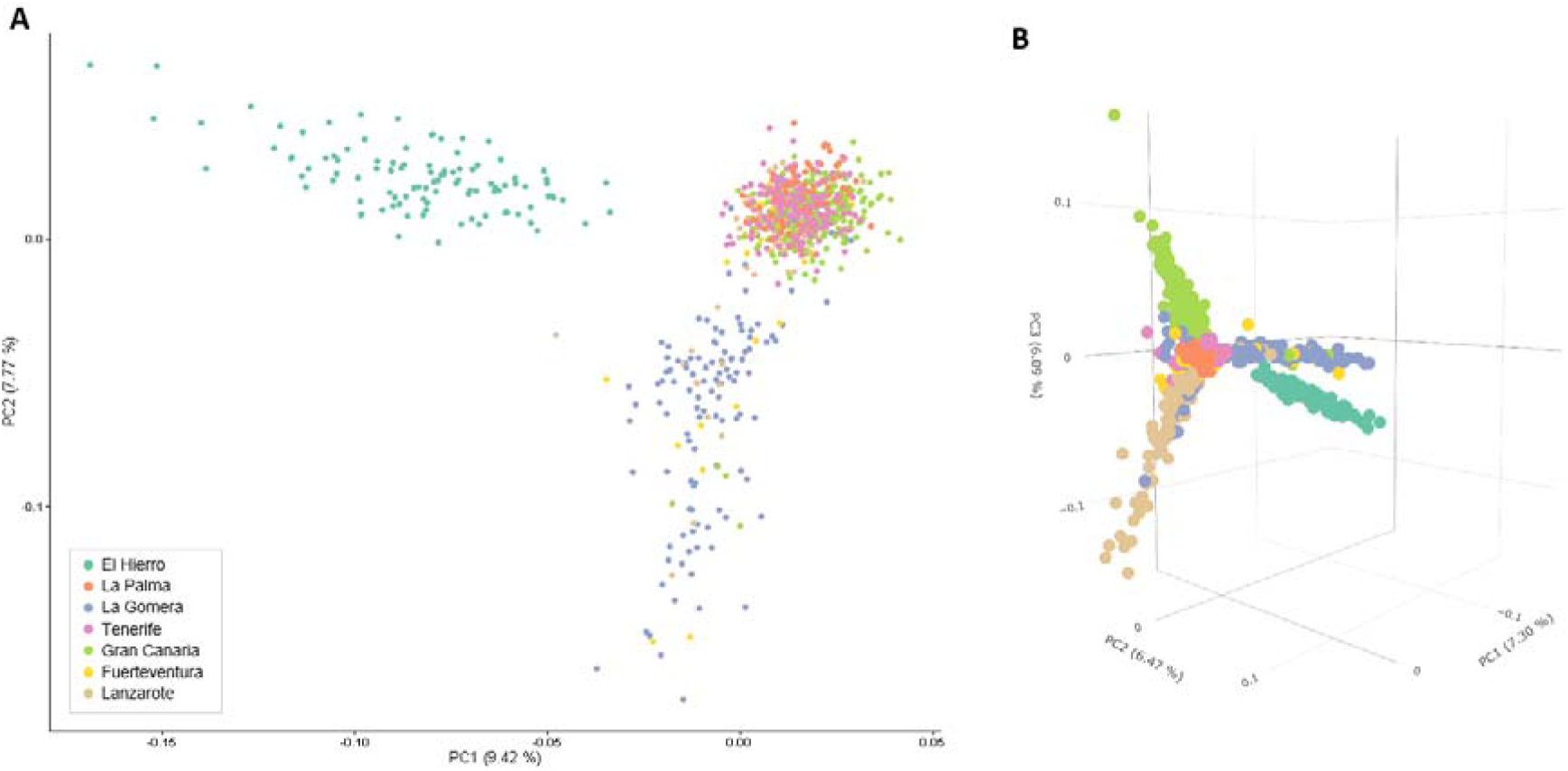
Representation of the first two (**A**) and the first three (**B**) principal components (PCs) of exome-wide genetic variation in Canary Islanders. **A**. Common variants (AAF >0.05) explaining 17.2% of variability. **B**. Rare variants (AAF <0.005), excluding singletons and doubletons, explaining 19.9% of variability.

To maximize discrimination among groups, we then relied on DAPC with all WES variants, revealing El Hierro as a clear outlier when compared to the other Canary Islands (**Figure 7**). El Hierro showed the largest number of variants with statistical differences in AAF among comparisons within Canary Islands in the LD1 (Fisher’s exact test after Bonferroni correction, *p*<2.86×10^-5^), except in the rare AAF range. We identified 15 variants with statistically different AAF between El Hierro and five of the other islands. These 15 variants were in *FLT4*, *MALRD1*, C1orf159, *PPME1*, C12orf42, LOC101929058, *TMTC1*, *ABHD17A*, *SLC6A3*, *CEP85L*, *MRPL20-AS1*, *ZBTB17*, *RNF220*, *CXCR4*, and *GRIP2*. Intriguingly, some of these genes have been associated with cardiovascular traits: *FLT4* associates with coronary heart disease (Kulminski et al. 2018), *ZBTB17* with dilated cardiomyopathy (Tadros et al. 2021), *MALRD1* with coronary artery disease in Type 1 diabetes (Charmet et al. 2018), and *RNF220* with heart rate (Eppinga et al. 2016; Verweij, van de Vegte, and van der Harst 2018). Additionally, *MALRD1*, *TMTC1*, *CEP85L*, and *RNF220* have been involved in diverse respiratory conditions (Lutz et al. 2015; Forno et al. 2017; Tian et al. 2017; Sakornsakolpat et al. 2019). In the rare AAF stratum, individuals from Gran Canaria clustered out when compared with the other populations (Supplementary Figure S3A). Within the Canary Islands, El Hierro and Gran Canaria exhibited two separate clusters (Supplementary Figure S3B).

**Figure 7.**
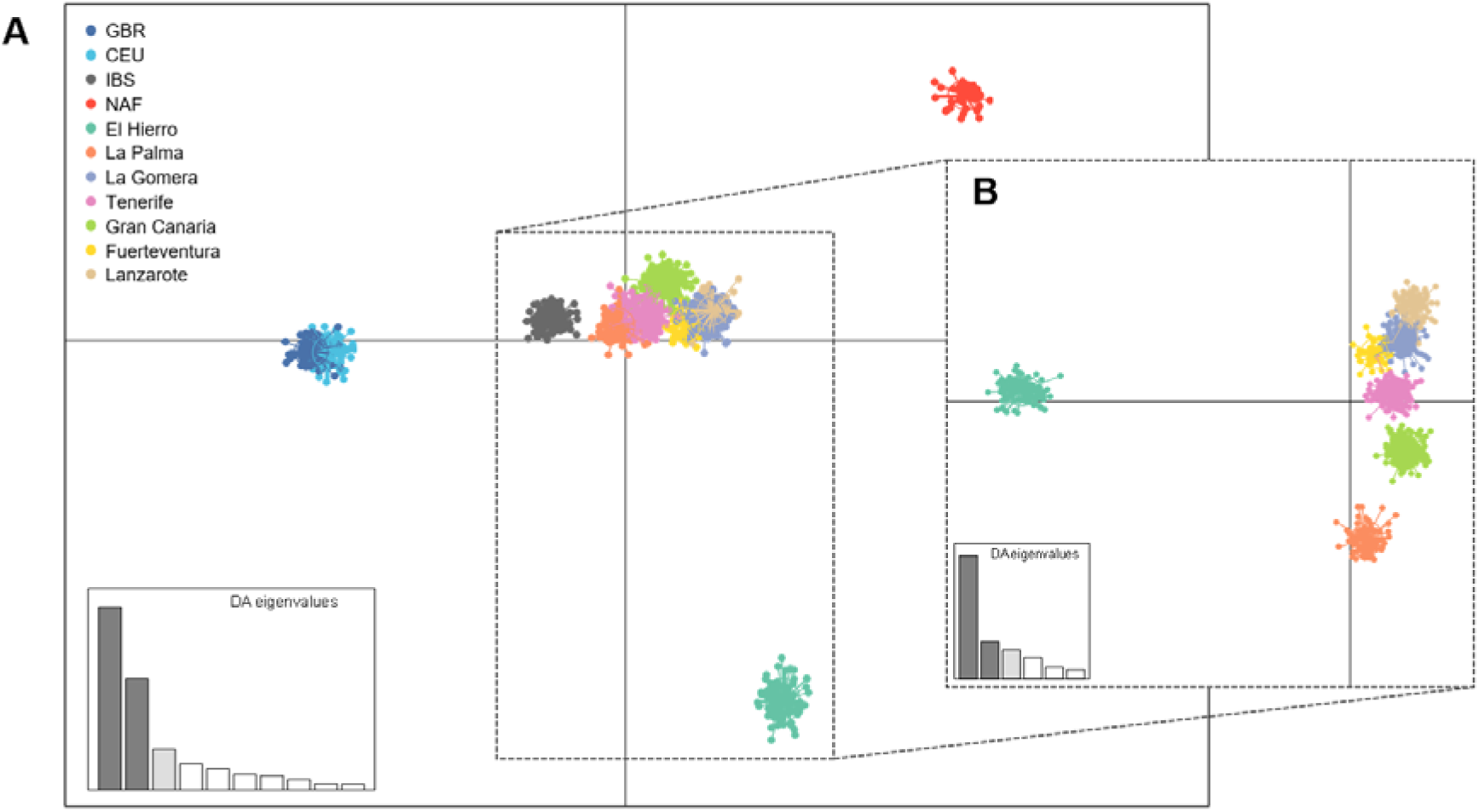
Discriminant Analysis of Principal Components (DAPC) for all variants. (**A**) Analysis including both Canary Islanders and reference populations (excluding outlier populations: SSA and FIN); and (**B**) Analysis only the Canary Islanders. LD1, horizontal axis; LD2, vertical axis. CEU, Utah Residents with Northern and Western European ancestry (1KGP); GBR, British (1KGP); IBS, Iberian population in Spain (1KGP); NAF, North African population.

### Genetic ancestry and ROHs in CIRdb

Genetic ancestry proportions in CIRdb individuals were previously obtained based on the SNP array data (Díaz-de Usera et al. 2022). Further details of CIRdb individuals are provided in Supplementary Table S11 and Supplementary Figure S4 evidencing an overall picture of 77.6 ± 3.71% (SD) of a genetic ancestry associated with a component prevailing in EUR, 18.9 ± 3.10% (SD) of a component prevailing in NAF, and 3.52 ± 1.51% (SD) of a component prevailing SSA. Based on local genetic ancestries, we previously identified genomic regions enriched in one of the ancestries and associated these regions with signals of selection (Guillen-Guio et al. 2018). By leveraging a sample that nearly doubled that of the previous study, we revealed previously identified regions and a novel one showing large deviation in the local genetic ancestry (**Figure 8**). The previously described regions included 2q21.3-2q22.1 (linked to lactase persistence) and 6p21.32 (in the HLA region), which were enriched in NAF and depleted in EUR ancestry, and 6p21.33, which was enriched in SSA ancestry. The novel region was located in 17q21.31 and was enriched in EUR and depleted in NAF ancestry (**Table 1**, Supplementary Material). This region overlaps with a well-known inversion site proposed to be under positive selection in European populations (Stefansson et al. 2005) and with pleiotropic effects across diseases including severe respiratory diseases, among others (Tantisira et al. 2008; Fingerlin et al. 2013; Noth et al. 2013; Degenhardt et al. 2022).

**Figure 8.**
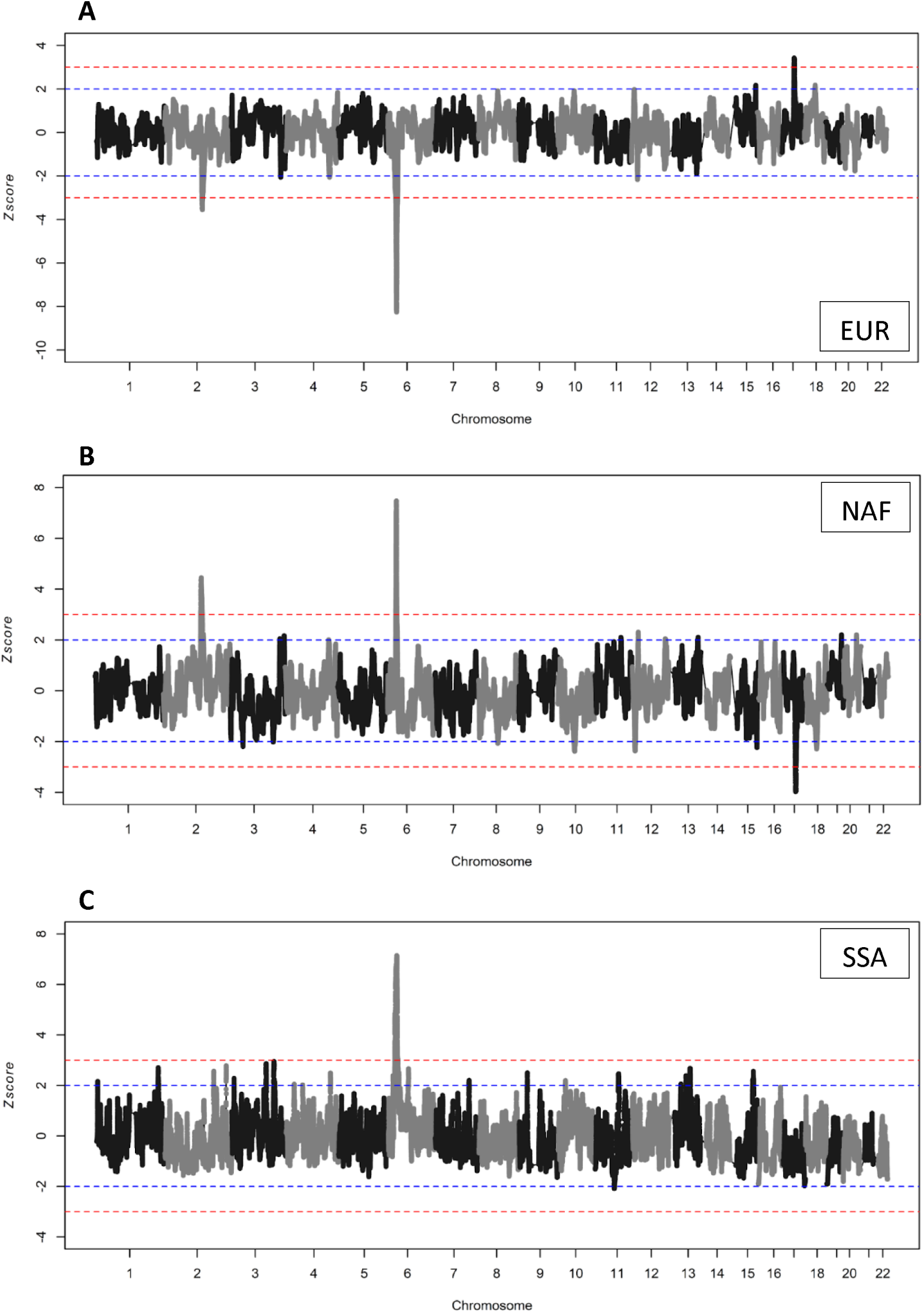
Genome-wide scan of deviations in local admixture for European (**A**), North African (**B**), and Sub-Saharan African (**C**) ancestry components in the current inhabitants of the Canary Islands from CIRdb. Horizontal dashed lines (blue>|2|; red>|3|) indicate z-score thresholds.

**Table 1.** Regions enriched or depleted in European (EUR), North African (NAF), and sub-Saharan African (SSA) local ancestries in Canary Islanders.

| Ancestry | Regions | Lead SNP | z-score | Mean local ancestry (%) | Genes |
| --- | --- | --- | --- | --- | --- |
| EUR | chr2: 135,338,640 - 137,893,387 | rs6430600 | -3.55 | 65.2 | *DARS1*,<br>*CXCR4* |
|  | chr6: 28,881,660 - 35,468,873 | (*) | -8.25 | 48.8 | *TSBP1* |
|  | chr17: 43,353,763 - 44,977,142 | rs1989480 | 3.42 | 89.6 | *LINC02210*-<br>*CRHR1* |
| NAF | chr2: 135,174,546 - 140,487,507 | rs6430600 | 4.45 | 32.0 | *DARS1*,<br>*CXCR4* |
|  | chr6: 30,074,163 - 33,956,165 | (**) | 7.47 | 40.8 | *TSBP1*,<br>*TSBP1-AS1* |
|  | chr17: 43,068,547 - 45,308,047 | rs1406072 | -3.97 | 7.20 | *KANSL1* |
| SSA | chr6: 23,934,383 - 35,337,197 | rs3130985 | 7.14 | 12.4 | *PSORS1C1* |

(\*) rs17208363, rs9368712, rs9368714, rs9380289, rs9296021, rs3132958, rs10484560, rs9405090, rs3129949, rs3132959, rs742582, rs1003878, rs910052, rs1076712, rs17840113, rs9366793, rs9348882, rs9357140, rs16870056, rs2022537
(\*\*) rs9268429, rs12528797, rs7758215, rs7757831, rs2894254, rs2395154, rs9268435, rs28895023

We used ROH estimates to assess isolation and inbreeding in the islands. In agreement with previous findings, El Hierro and La Gomera showed the highest estimates for the average total ROH, the total ROH length (Mann-Whitney U-test, *p*<1×10^-3^ for all pairwise comparisons), and the number of fragments in ROHs (Mann-Whitney U-test, *p*<1×10^-5^ for all pairwise comparisons) compared to all other islands (**Figure 9**, Supplementary Figure S5 and S6, Supplementary Table S12).

**Figure 9.**
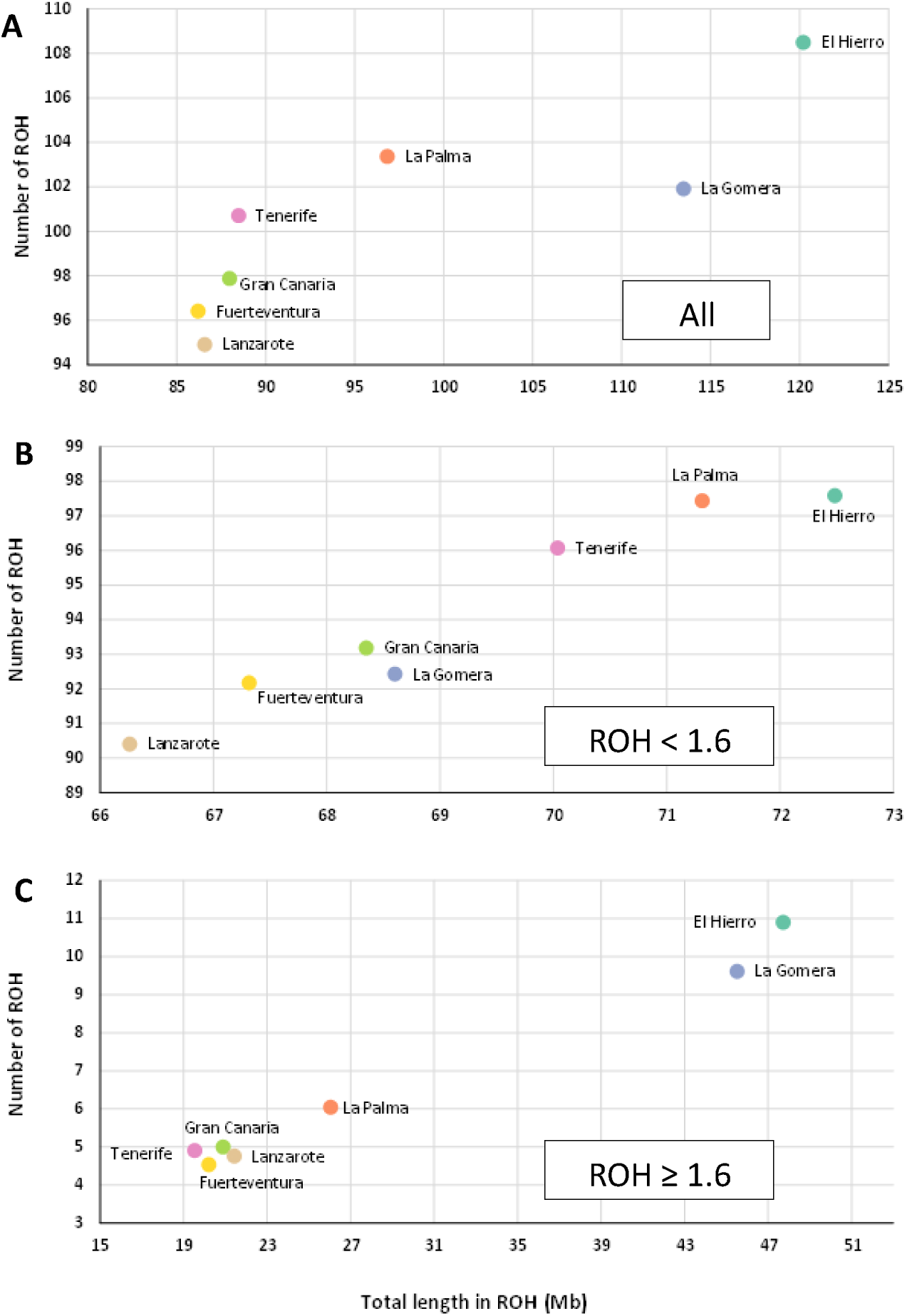
Comparison of the average total ROH length (Mb) and the number of ROH fragment per island. Top (**A**), all ROHs; Middle (**B**), ROHs <1.6 Mb; Bottom (**C**), ROHs ≥1.6 Mb.

### A scan for selective sweep around PRUNE1 and a PheWAS of the exonic variant

Among the prioritized common variants in DAPC LD1, we identified 1,407 WES variants that showed no significant AAF differences between the Canary Islanders (altogether or for each island by separate) and the NAF population (Fisher’s exact test, Bonferroni-corrected *p*>3.33×10^-5^). Six out of those variants showed statistically significant differences in AAF between the Canary Islands and EUR populations (Fisher’s exact test, Bonferroni-corrected *p*<3.33×10^-5^) despite being frequent variants (AAF>10%) in the Canary Islands (**Table 2**, Supplementary Table S13). Of those, we focused on rs3738476 in chromosome 1, a variant exonic to Prune Exopolyphosphatase 1 (*PRUNE1*) gene that showed the largest difference in AAF between the Canary Islands (87.2%) and NAF (91.2%) compared to EUR populations (AAF range: 49.5% and 52.7%), as quantified by the PBS genetic distance (0.303) and the pairwise F_ST_ distances between NAF or Canary Islanders *vs.* EUR (Supplementary Tables S14 and S15, respectively). To evaluate the existence of a putative selective sweep in the region, we conducted an iSAFE scan based on WGS data in a subset of the participants and in IBS and NAF in the surrounding region of this variant (chr1:150,520,898-151,498,624) (**Figure 10**). The region with the highest iSAFE score was chr1:151,010,521-151,116,279 (Canary Islands: iSAFE score >0.12, peak at 0.131; NAF: iSAFE score >0.11, peak at 0.136), while the signal tempered for IBS. This region harbors other 36 variants associated with body mass index (BMI)-related traits and functionally linked to nine genes involved in cardiovascular traits (*BNIPL*, C1orf56, *CDC42SE1*, *CTSS*, *LYSMD1*, *MLLT11*, *PRUNE1*, *SEM6C*, and *TNFAIP8L2*) (Pulit et al. 2019) and diabetes mellitus (*BNIPL*, C1orf56, *CDC42SE1*, *CTSS*, *MLLT11*, and *TNFAIP8L2*) (Vujkovic et al. 2020) (Supplementary Table S16). We then conducted a PheWAS to examine possible associations between the rs3738476 variant at *PRUNE1* and the 66 available phecodes of participants (**Figure 11**). Although no associations were evidenced at statistical significance, nominally significant associations (*p*<0.05) were found with respiratory disease, intestinal diverticula, and stomach cancer.

**Table 2.** Variants with similar alternative allele frequencies in Canary Islanders and the North African populations but differing from European populations.

| **Chromosome** | 1 | 2 | 5 | 8 | 14 | 15 |
| --- | --- | --- | --- | --- | --- | --- |
| **Position (hg19)** | 151,006,539 | 135,911,422 | 33,963,870 | 19,816,934 | 102,349,907 | 28,510,895 |
| **Reference allele** | C | T | C | T | A | G |
| **Alternative allele** | A | C | T | C | G | A |
| **Location** | Exonic | Exonic | Exonic | Intronic | Intronic | Intronic |
| **rsID** | rs3738476<sup>†</sup> | rs17261772 | rs26722<sup>†</sup> | rs301<sup>†</sup> | rs2720207 | rs2240202 |
| **Gene** | *PRUNE1* | *RAB3GAP1*<sup>†</sup> | *SLC45A2* | *LPL* | *PPP2R5C*<sup>†</sup> | *HERC2*<sup>†</sup> |
| **Associated trait** | Body mass index | Body fat percentage; LDL and total cholesterol levels; obesity | Cutaneous melanoma, hair color, facial wrinkles (forehead) | Diacylglycerol levels, metabolic syndrome | Protein levels | Cutaneous melanoma, hair, and eye color |
| **AAF** |  |  |  |  |  |  |
| **Canary Islands** | 0.872 | 0.557 | 0.114 | 0.237 | 0.313 | 0.142 |
| **CEU** | 0.495* | 0.217* | 0* | 0.424 | 0.192 | 0.066 |
| **GBR** | 0.527* | 0.275* | 0.005 | 0.522* | 0.104* | 0.005* |
| **IBS** | 0.519* | 0.444 | 0.079 | 0.458* | 0.201 | 0.187 |
| **NAF** | 0.913 | 0.558 | 0.173 | 0.163 | 0.317 | 0.192 |

AAF, alternative allele frequency; Alt, alternative allele; CEU, Utah Residents with Northern and Western European ancestry (1KGP); Chr, chromosome; GBR, British from England and Scotland (1KGP); IBS, Iberian population in Spain (1KGP); NAF, North African population; Ref, reference allele. \*Statistical differences with all Canary Islands populations after Bonferroni correction ( $p < 3.33 \times 10^{-5}$ for Fisher's exact test). <sup>†</sup>Variant or gene associated with the trait.

**Figure 10.**
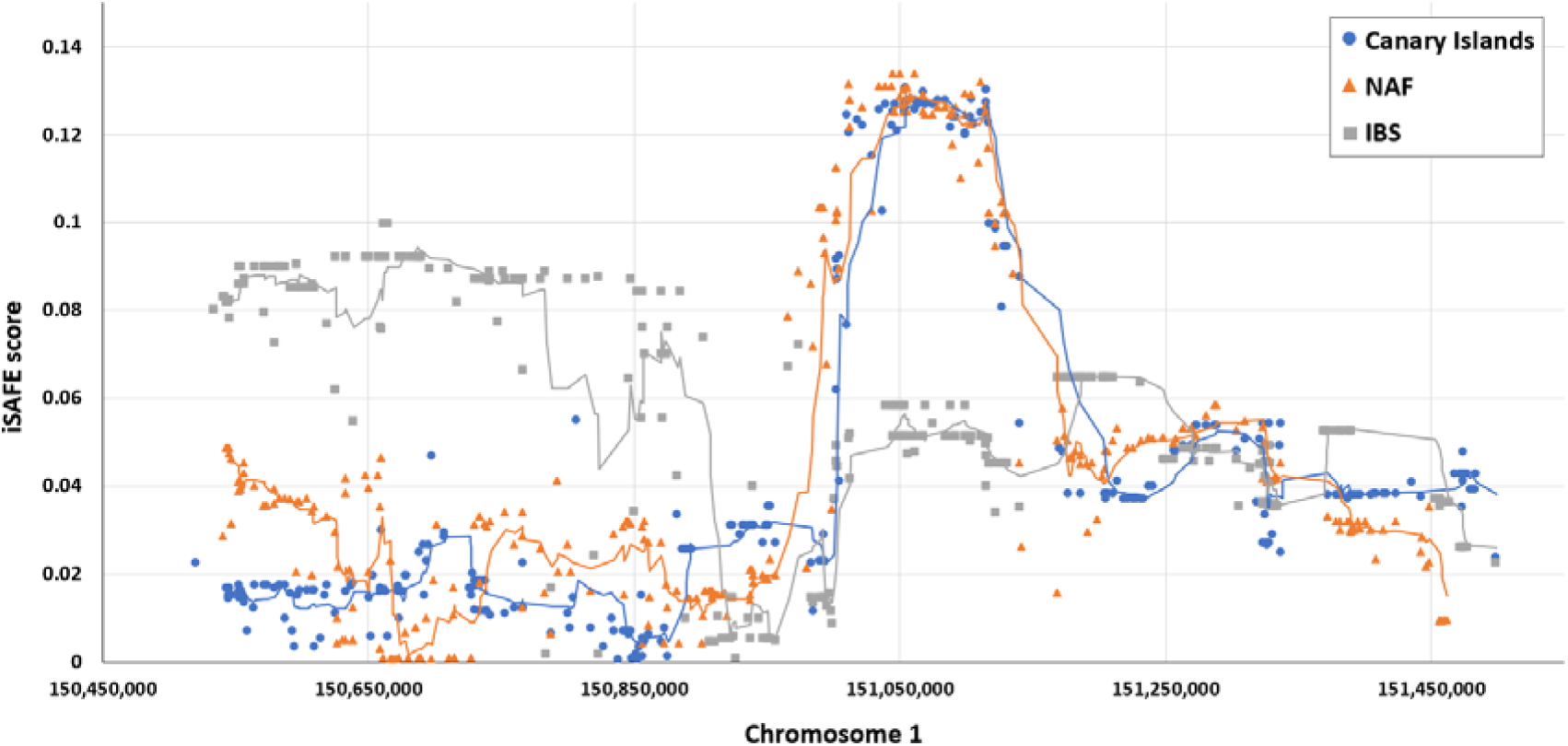
iSAFE scan of whole-genome sequencing data in the genomic region surrounding rs3738476. Lines represent the moving average for each population. IBS, Iberian population in Spain (1KGP); NAF, North African population.

**Figure 11.**
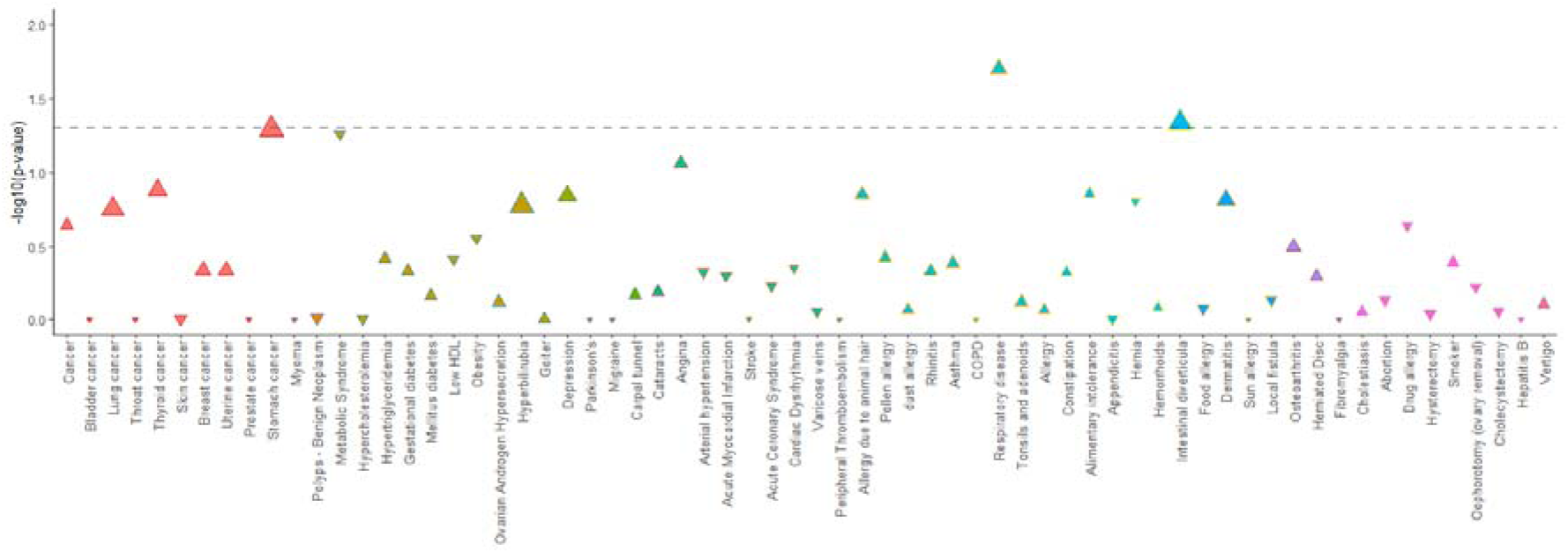
Results from the logistic regression analysis testing the association between the rs3738476 variant and the incidence of 66 phecodes, with age and sex included as covariates. Each triangle represents the *p*-value of the association, pointing upward or downward depending on whether the variant is a risk or a protective factor. The size of the triangle corresponds to the odds ratio (OR), and its color indicates the respective ICD-9 category. The horizontal gray line represents the nominal significance threshold (*p*=0.05).

## Discussion

We developed CIRdb as the first exome-wide catalog of the natural genetic variation in the current inhabitants of the Canary Islands, the European population with the highest fraction of NAF ancestry reported to date. We found that approximately 15.1% of the variants found were novel at the time the analyses were conducted, emphasizing the key necessity for developing a population-specific catalog of genetic diversity in this archipelago (Díaz-de Usera et al. 2022). A similar scenario was observed in other populations like Cilento in South Italy (Nutile et al. 2019) and the Iranian population (Fattahi et al. 2019). These findings highlight the importance of genetic studies in diverse populations and the potential for unique discoveries. Strikingly, the island of La Palma exhibited the highest average of novel variants per individual, making it a compelling candidate for future studies on disease-causing variation. Our diverse analyses combining WES, SNP array, and WGS data comparing the genetic background of the current Canary Islanders with other populations from Europe and Africa reinforced the recent admixed nature of the population and the footprints of population isolation, which were most salient in two of the smallest islands (El Hierro and La Gomera). This resource allowed us to identify genetic variation that could have significant biomedical implications for prevalent diseases in the Canary Islands, including metabolic, infectious, and respiratory diseases, among others, and in skin pigmentation and sunburn. We also identified a new region showing large deviations in local ancestry around a common inversion (17q21.31) known to be under positive selection in European populations and showing pleiotropic effects in human diseases. Our analyses also prioritized the region around *PRUNE1* to be under putative positive selection in Canary Islanders, a genomic region associated with body mass index (BMI), and metabolic and cardiovascular disorders. We expect that the CIRdb exome-wide catalog of the natural genetic variation in the Canary Islands will constitute a key resource to assist in genetic diagnosis of patients and in the identification of disease risks, as described elsewhere (Lorente-Arencibia et al. 2022).

The inhabitants of the Canary Islands have had a unique history of admixture and isolation in the southwestern European context. Although genetic isolation footprints have been well-documented, at least for some of the islands such as El Hierro (Ordóñez et al. 2017; Guillen-Guio et al. 2018), its impact on the prevalence of genetic diseases in the population remains largely unexplored. Many studies support a higher prevalence of cardiovascular (Cabrera de León et al. 2006; Bueno, Hernáez, and Hernández 2008; Marcelino-Rodríguez et al. 2016; Rodríguez-Esparragón et al. 2017) and respiratory and allergic diseases (Sánchez-Lerma et al. 2009; Juliá-Serdá et al. 2011) in the Canary Islands compared to mainland regions of Spain. Some monogenic disorders have been linked to founder effects in particular islands, such as Fanconi anemia in La Palma (Castella et al. 2011), type 1 primary hyperoxaluria in La Gomera (Santana et al. 2003), and Wilson disease in Gran Canaria (García-Villarreal et al. 2000; Lorente-Arencibia et al. 2022). This evidence is in line with our findings and the patterns of genetic differentiation in the exonic regions and the presence of private variants in current Canary Islanders.

There is support for the biomedical implications of the recent North African genetic influences in the European population context (Botigué et al. 2013). Accordingly, we prioritized common genetic variants -with AAF shared between Canary Islanders and North Africans while diverged in Europeans, including Iberians- that were associated with risk for cardiovascular disorders. Some of these were rs301 in *LPL* that is associated with insulin resistance and atherosclerosis (Deo et al. 2009; Kraja et al. 2011) and rs17261772 in *RAB3GAP1* that is associated with sudden cardiac arrest (Huertas-Vazquez et al. 2013). We also prioritized genomic regions that showed evidence of a selective sweep in the Canary Islanders and North Africans but not in Iberians, with a peak at a synonymous exonic variant (rs3738476) in *PRUNE1*. The risk allele rs3738476_A, which is near fixation in the Canary Islanders and North Africans, was previously associated with BMI-related traits (Pulit et al. 2019). The available evidence supports that the risk variant could affect gene function by creating a new “GGACU” sequence that would be susceptible to being methylated (Liu et al. 2020). A PheWAS of this variant in the Canary Islanders also provided tentative links with other diseases, including respiratory disease, intestinal diverticula, and stomach cancer, although these results should be interpreted with caution given that statistical significance was not reached.

The following constitute the main limitations of the study. First, whole-exome sequencing was based on short-read technology applied to nearly 1,000 unrelated individuals reporting at least two ancestors born in the archipelago. This limited our ability to study ultra-rare variants and the optimal assessment of structural variation, which also pose significant risks for disease (Huddleston and Eichler 2016). Second, sequence variation at non-exonic regions remains largely uncovered, which will necessitate whole-genome approaches to better understand disease architecture and provide further insights into population history (Collins et al. 2020). Third, since the study prioritized individuals without history of severe diseases as well as no cardiovascular, metabolic, immunologic, or cancer conditions, the PheWAS was based on a limited number of samples that were positively ascertained for the traits assessed. Fourth, given that we used GRCh37/hg19 as the reference, our study may have missed information from important genomic regions that could be assessed by leveraging the most recent version of the human reference genome, i.e., T2T-CHM13 (Nurk et al. 2022). Fifth, since genetic sequence variation in North Africans is central to understanding the genetic diversity in the Canary Islanders, some of our results may be affected by the limited sample size of North Africans used as a reference due to their underrepresentation in global genetic studies (Gurdasani et al. 2015; Serradell et al. 2024).

## Conclusion

We present CIRdb, the first catalog of genetic variation obtained by whole-exome sequencing in current inhabitants of the Canary Islands. We found evident patterns of isolation in El Hierro and La Gomera. We also identified two genetic loci with well-known links with complex disease risks evidencing patterns indicative of selective processes and shared patterns of genetic differentiation between North Africans and Canary Islanders that are distinct from Europeans.

## Data Availability

Data supporting the findings are available as part of the manuscript or from the supplementary files. Access to the raw sequence dataset is restricted to qualified researchers under institutional agreements. Data access requests must be reviewed before release.

## Acknowledgements

We would like to thank the support from our colleagues from the Teide-HPC Supercomputing facility (http://teidehpc.iter.es/en), which was funded by INP-2011-0063-PCT-430000-ACT (INNPLANTA program) from the Spanish Ministry of Economy and Competitiveness.

## Author contributions

Conceptualization: C.F.; Methodology: A.D.-d.U., R.C.-G., J.M.L.-S., and C.F.; Investigation: A.D.-d.U., B.G.-G., I.M.-R., A.Co., R.G.-M., and A.Ca.; Formal Analysis: A.D.-d.U., L.A.R.-R., A.M.-B, J.M.L.-S., D.J., I.M.-R., and R.C.-G.; Data Curation: A.D.-d.U., L.A.R.-R., A.M.-B, R.G.-M., and D.J.; Resources: M.C.R.-P., A.C.-d.-L., A.C.; Supervision: C.F.; Funding Acquisition: C.F.; Writing – Original Draft Preparation: A.D.-d.U. and C.F.; Writing – Review & Editing: all authors.

