## Supplementary material for "Expanding CIRdb, a comprehensive catalog of whole-exome sequencing data of Canary Islanders"

**Supplementary Methods**

Sample selection and SNP array genotyping

DNA was extracted from peripheral blood using the Blood genomicPrep Mini Spin Kit (Cytiva, Marlborough, MA, USA) following the manufacturer’s recommendations. Prior to the sequencing process, genotyping was performed for the 587,352 variants contained in the Axiom® Genome-Wide Human CEU 1 Array (Affymetrix, Santa Clara, CA) supported by the National Genotyping Center (CeGen), Universidad de Santiago de Compostela Node. Image files were processed, and the genotyping quality controls (QCs) were conducted using the AffyPipe v2.10.0 open-source pipeline (Nicolazzi, Iamartino, and Williams 2014) followed by additional standard QCs applied on samples using the R 3.2.2 environment and PLINK v1.9 as described elsewhere (Guillen-Guio et al. 2018). Samples with sex discordances (as recorded in medical records versus genetically inferred), genotype call rate <0.95, outlier heterozygosity rate (average ± [4 x SD]), and kinship relatedness (PIHAT>0.2) were removed. We also filtered out non-autosomal variants, variants strongly deviating from Hardy-Weinberg equilibrium (*p*<1x10^-6^), variants with a call rate <0.95, and variants with a minor allele frequency below 1% (MAF <0.01).

Reference population datasets

The North African population dataset (NAF) encompassed whole-genome data of 32 individuals from distinct countries gathered for a previous study sequenced with Illumina HiSeq 2000 using PE100 reads or HiSeq 4000 using PE150 reads (Serradell et al. 2024). Before merging with EUR and SSA references, several filters were used on the NAF dataset: keeping sites with callable regions and read depth (DP) ≥10, flagged as PASS in the FILTER field, and Genotype Quality (GQ) ≥20. Additionally, multi-nucleotide variants (MNP) were converted to single-nucleotide variants (SNPs). Biallelic variants from the NAF dataset were merged with EUR and SSA populations using the *merge* command from BCFtools (Danecek et al. 2021), considering the identifier as CHR:POS:REF:ALT.

Whole-exome sequencing: library quantification

Libraries were checked using the Qubit^TM^ dsDNA HS Assay Kit on a Qubit® 3.0 Fluorometer (Thermo Fisher Scientific, Waltham, MA, USA) and the Agilent D1000 and High Sensitivity D1000 ScreenTape Assays on a 4200 TapeStation system (Agilent Technologies, Santa Clara, CA, USA).

Whole-exome sequencing: processing of GVCF

Variant calling per sample was carried out with the GATK HaplotypeCaller and the output was provided in GVCF (Genomic Variant Call Format) mode. GVCF files were used as input for GATK GenotypeGVCFs obtaining one multisample genotyped raw VCF file. This raw VCF file was recalibrated with GATK Variant Quality Score Recalibration (VQSR) to reduce false positives and to reach a high level of sensitivity. The resulting recalibrated-multisample VCF from the VQSR step was split per-sample and used to refine the initial GVCF files, producing a refined-recalibrated final set which contained both refined-recalibrated-variant and non-variant sites in genomic VCF format (GVCF). Based on the splitted VCF data, non-variants sites in the GVCFs were refined per sample following next steps: a) if DP<10, a missing genotype (GT=./.) was assigned; b) if DP ≥10 and genotype did not include at least one alternative allele (GT=./. or GT=0/0), genotype remained as detected by the method; c) if DP≥10 and genotype included at least one alternative allele, QUAL score normalized using Allele Depth (QD) was evaluated. Therefore, missing genotype (GT=./.) was assigned for QD<2 and homozygous reference (GT=0/0) was established as genotype for QD>2. These criteria were applied in sites corresponding to variants initially detected in the GVCF but not finally called in the joint-genotyped VCF.

Whole-exome sequencing: post-variant calling filters

The following filters were applied after variant calling:

1. Selection of sites located in the manifest file of target regions provided by the commercial target-capture solution;
2. Choice of callable regions;
3. Selection of sites with DP≥10;
4. Removal of mitochondrial chromosome; and
5. Filtering out of sites without a PASS flag and/or GQ<20.

Using BCFtools v1.10 (Danecek et al. 2021), individual GVCFs were merged into a multisample GVCF which was subjected to additional filters, including the conversion of multi-nucleotide variation into single-nucleotide variation and the selection of variant sites. Furthermore, variants susceptible to systematic sequencing bias in whole-genome analysis were removed (Freeman et al. 2020). Variants with a low call rate (<0.80), significant deviation from Hardy-Weinberg equilibrium (*p*<1x10^-6^), and with negative values for Inbreeding Coefficient (<-0.3), based on gnomAD v2.1 project (Karczewski et al. 2020), were also excluded. Previously to the establishment of the final catalog of variants, a normalization step by means of BCFtools v1.10 was used to avoid discrepancies between variant callers and annotation software.

Whole-exome sequencing summary

Whole-exome sequencing of the CIRdb cohort provided an average of 8.41 Gbases ± 2.15 standard deviation (SD), 99.7 ± 0.40% (SD) of which were mapped against the reference genome and passed filters. The average on-target depth of coverage was 51.9 ± 19.6X (SD) (range: 15.7-163X). The average on-target breadth of coverage was 97.4 ± 1.09% (SD) at >1X, and 60.8 ± 13.8% (SD) at >30X.

Selective sweep analysis

Variants from whole-genome sequence data from 32 NAF, 66 Canary Islands, 58 IBS from 1KGP, and 10 YRI individuals randomly selected within 107 from 1KGP (acting as outgroup population) were assessed for the selective sweep analysis. For this purpose, the Canary Islanders whole-genome data was obtained with HiSeq 4000 using PE150 reads for a subset of the donors. Read alignment was carried out using the Burrows-Wheeler Aligner with Maximal Exact Matches algorithm (BWA-MEM) v0.7.12-r1039 (Li and Durbin 2009) and GRCh37/hg19 was used as the reference genome. The variant calling was performed using an in-house bioinformatics pipeline following the Genome Analysis Toolkit (GATK) v4 Best Practices guidelines (O’Connor and van der Auwera 2020). Biallelic SNPs flagged with PASS in the FILTER field were phased within the EagleImp-Web service (Wienbrandt, Lars, Christoph Prieß, Jan Christian Kässens, Andre Franke, Franziska Uhing, and David Ellinghaus 2022).

**Supplementary Results**

Missense variants

A high proportion of variation was identified as missense variants (41.3%, moderate impact), meaning that there exists a prediction of an amino acid change in the final protein which may alter its function. There were no statistically significant differences among the number of missense variants among the islands (one-way ANOVA test, *p*=0.562). The second and the third most numerous variants, based on consequence classification, were synonymous and intron variants representing 25.5% and 14.1%, respectively. The rest of variants (19.2%) were distributed in 22 different groups. From a total of 159,940 missense variants, 21,001 were novel (i.e., not declared in dbSNP154, 1KGP, gnomAD exome, TOPMed v5, or HGDP-CEPH), with more than 98.4% classified as rare, including 14,520 singletons, and 3,589 doubletons. La Palma provided the highest average of missense novel variants per individual, being significantly different from El Hierro and Fuerteventura (one-way ANOVA test, *p*=0.045). Missense novel variants provided the following pathogenicity distribution based on InterVar annotation: 434 likely benign, 209 likely pathogenic, one likely pathogenic + VUS, one pathogenic variant, and 20,356 VUS. Applying the M-CAP score, 43.7% of VUS declared by InterVar could be cataloged such as possibly pathogenic variants based on threshold above 0.025.

Loss-of-function variants

Additionally, we performed an assessment of loss-of-function (LoF) variants. Out of the 387,555 total variants, 16,542 were labeled as high confidence (HC) by LOFTEE v1.0, of which 1,252 provided C-score values above MSC (99% CI) and pLI values above 0.9. No differences in the presence of LoF variants were observed among islands (ANOVA test, *p*=0.373). HC LoF variants with C-score value above MSC (99% CI) and pLI >0.9 were distributed on 776 genes, where a total of 123 genes encompassed variants above the mean (i.e., two variants per gene). According to the first 20 biological processes identified by GO analysis, enrichment in these 123 genes supported links to aorta development, regulation of catalytic and lipid kinase activity, and aortic valve and ventricular compact myocardium morphogenesis, among others (**Supplementary** **Figure S4**, **Supplementary Table S5**).

Prioritization analysis from the DAPC LD1

As observed for rs3738476 in *PRUNE1*, only a few variants for CEU (2), GBR (4), and IBS (1) populations, showed clear differences in AAF compared to the Canary Islands data considered in aggregate (**Table 2**). The exonic variant in *RAB3GAP1* gene (rs17261772) was related to traits which have a close relationship with cardiovascular and respiratory systems, such as body fat percentage, LDL and total cholesterol levels or obesity, and asthma. Additionally, this variant has been established as a protein quantitative trait locus (pQTL) and/or expression quantitative trait locus (eQTL) of some genes implicated in lactose metabolism (i.e., *LCT, MCM6,* and *DARS1* genes). Variants in chromosomes 5 (rs26722) and 15 (rs2240202) are involved in skin, hair, and eye pigmentation, with rs2240202 acting as an eQTL of *HERC2* and *OCA2,* both genes in line with the same traits. The intronic variant in *LPL* gene (rs301) plays a main role in lipoprotein metabolism as well as in some complications derived from it such as coronary artery or heart diseases.

Local ancestry blocks

The EUR-related region in chr2 displayed an average local ancestry of 65.2% for its lead SNP which was an intergenic variant located between the *DARS1* and *CXCR4* genes (2q21.3 and 2q22.1), whereas the regions in chr6 (with all lead SNPs located within *TSBP1*, 6p21.32, including two exonic variants and 18 intronic) and in chr17 (lead SNP located in an intronic region of the *LINC02210-CRHR1* gene, 17q21.31) exhibited an average local ancestry of 48.8% and 89.6%, respectively (**Table 2**). The loci with clear deviation in NAF ancestry covered regions from chr2 (2q21.3 and 2q22.1) (with the same lead SNP that from EUR-related peak, with an average local ancestry of 32.0%), chr6 (showing all lead SNPs as intergenic variants flanked by *TSBP1* and *TSBP1-AS1*, 6p21.32, with an average local ancestry of 40.8%), and chr17 (providing the lead SNP with an average local ancestry of 7.20% and within the *KANSL1* gene, 17q21.31). The locus enriched in SSA ancestry was located in *PSORS1C1* gene in chr6 (6p21.33), where the intronic lead SNP provided an average local ancestry of 12.4%.

The lead SNP from chr2 (rs6430600) has been related to *LCT* and *MCM6* genes by means of the Variant to Gene (V2G) score (Ghoussaini et al. 2021), being involved in metabolism of the lactose. Lactase persistence, one of the strongest examples of positive selection (Ségurel and Bon 2017) on the human genome, is a heterogeneous trait whose worldwide distribution range from 5% to almost 100%. Despite the interest aroused by this trait, nowadays there are populations that still are underrepresented in this context (i.e., North African population). Thus, the signal observed in our study may pinpoint a positive selection in this region caused by North African genomic ancestry. Strikingly, two of the prioritized variants from DAPC, for both scenarios considered, were located in this NAF-enrichment region in chr2 (rs61731939 and rs17261772). When regions in chr6 were considered, different variants acted as eQTLs of five of the eight classical HLA alleles (*HLA-B*, *HLA-C*, *HLA-DBQ1*, *HLA-DRB1*, *HLA-DQA1*) and of several non-classical HLA alleles. The human leukocyte antigen genes are located in the major histocompatibility complex (MHC) which is the most variable region in the human genome, characterized by a high degree of polymorphism and numerous functional genes. Studies around these genomic coordinates have emphasized an important selective process, not only in a balancing scenario such as could be observed in European and African populations (Bitarello et al. 2018; DeGiorgio, Lohmueller, and Nielsen 2014), but also in positive selection for African populations (de Bakker et al. 2006) and in negative selection in a population from North America (Lindo et al. 2016). Therefore, peaks in chr2 and chr6 signals may be explained as the result of putative selective processes in these regions within the Canary Islanders genome.

Lindo, John, Emilia Huerta-Sánchez, Shigeki Nakagome, Morten Rasmussen, Barbara Petzelt, Joycelynn Mitchell, Jerome S. Cybulski, Eske Willerslev, Michael DeGiorgio, and Ripan S. Malhi. 2016. “A Time Transect of Exomes from a Native American Population before and after European Contact.” *Nature Communications* 7 (November): 13175.

Nicolazzi, Ezequiel L., Daniela Iamartino, and John L. Williams. 2014. “AffyPipe: An Open-Source Pipeline for Affymetrix Axiom Genotyping Workflow.” *Bioinformatics* 30 (21): 3118–19.

O’Connor, Brian D., and Geraldine van der Auwera. 2020. *Genomics Analysis with Spark, Docker and Clouds*. Sebastopol, CA: O’Reilly Media.

Ségurel, Laure, and Céline Bon. 2017. “On the Evolution of Lactase Persistence in Humans.” *Annual Review of Genomics and Human Genetics* 18 (August): 297–319.

Serradell, Jose M., Jose M. Lorenzo-Salazar, Carlos Flores, Oscar Lao, and David Comas. 2024. “Modelling the Demographic History of Human North African Genomes Points to a Recent Soft Split Divergence between Populations.” *Genome Biology* 25 (1): 201.

Wienbrandt, Lars, Christoph Prieß, Jan Christian Kässens, Andre Franke, Franziska Uhing, and David Ellinghaus. 2022. “EagleImp-Web: A Fast and Secure Genotype Phasing and Imputation Web Service Using Field-Programmable Gate Arrays.” *BioRxiv*, 2022.02.24.481790.

**Supplementary Tables**

| **Supplementary Table S1.** Software, databases, and plugins used for variant annotation. | | | |
| --- | --- | --- | --- |
| **Software** | **Database / Plugin** | **Release / Version** | **Annotation information** |
| ANNOVAR | RefGene | r20190929 | Genes |
|  | cytoBand | r20180813 | Regions |
|  | ClinVar | r20210501 | Phenotypes |
|  | dbnsfp | v4.1a | rsID number |
|  | gnomAD exome | v2.1.1 | Allele frequency in other populations |
|  | 1KGP | r2015aug | Allele frequency in diverse populations |
| The Ensembl VEP | dbSNP human | Build 154 | rsID number |
|  | Combined Annotation Dependent Depletion (CADD) | v1.6 | Pathogenicity prediction score |
|  | Loss-Of-Function Transcript Effect Estimator (LOFTEE) | v1.0 | Pathogenicity prediction score |
|  | Probability of a gene being loss-of-function intolerant (pLI) | r0.3 | Pathogenicity prediction score |
| InterVar | - | - | Clinical interpretation of genetic variants based on the 2015 ACMG-AMP Guidelines (Richards et al. 2015) |
| BCFtools | Mendelian Clinically Applicable Pathogenicity (M-CAP) | v1.4 | Pathogenicity likelihood score |
|  | Mutation Significance Cutoff (MSC) | v1.6 | Gene-level and gene-specific phenotypic impact cutoff values |
|  | Online Mendelian Inheritance in Man (OMIM) | r20210128 | Relationships between human genes and phenotypes |

| **Supplementary Table S2**. Total number of variants per island and average per island, per individual. SD, standard deviation. | | | |
| --- | --- | --- | --- |
| **Island** | **Samples** | **Variants per island** | **Average of variants per island per individual (± SD)** |
| El Hierro | 106 | 120,273 | 25,729 ± 815 |
| La Palma | 99 | 143,035 | 25,413 ± 1,143 |
| La Gomera | 145 | 163,321 | 25,601 ± 1,709 |
| Tenerife | 165 | 184,372 | 25,632 ± 678 |
| Gran Canaria | 215 | 208,919 | 25,714 ± 1,488 |
| Fuerteventura | 47 | 111,100 | 25,664 ± 842 |
| Lanzarote | 114 | 152,656 | 25,478 ± 2,257 |
| NIA | 29 | 94,131 | - |

**Supplementary Table S3.** Association between the 16 variants that showed statistically significant differences in alternative allele frequencies within island populations and the TOP5 V2G genes from Open Targets Genetics and the TOP5 diseases/phenotypes associated from Open Target Platform. Alt, alternative allele; Ref, reference allele. ^a^From GRCh37/hg19 coordinates; ^b^Most severe consequence; ^c^V2G score: Variant to Gene score; ^d^From Open Targets Genetics; ^e^From Open Targets Platform; ^f^El Hierro showed significant differences in the alternative allele frequency compared to these islands. El Hierro always showed higher alternative allele frequency values. Available as a separate xls file.

**Supplementary Table S4**. Enrichment analysis by means of Enrichr. Available as a separate xls file.

**Supplementary Table S5**. GeneSCF annotation in the 123 genes which encompassed variants above the mean (> two variants) considering HC LoF variants with C-score > MSC and pLI >0.9. Available as a separate xls file.

**Supplementary Table S6**. Putative pathogenic novel variants. Available as a separate xls file.

**Supplementary Table S7.** Studies from GWAS catalog showing associations with the genes linked to putative pathogenic variants. Available as a separate xls file.

| **Supplementary Table S8.** Distribution of consequence considering total, not shared, non-exclusively shared, and exclusively shared variants with North African population. Consequences were extracted from Ensembl VEP. | | | | | |
| --- | --- | --- | --- | --- | --- |
| **Consequence** | **Total** | **Not shared with NAF** | **Shared with NAF** | **Non-exclusively shared with NAF** | **Exclusively shared with NAF** |
| Splice acceptor variant | 4,771 | 4,603 | 168 | 131 | 37 |
| Splice donor variant | 1,428 | 1,227 | 201 | 170 | 31 |
| Stop gained | 4,313 | 3,806 | 507 | 418 | 89 |
| Frameshift variants | 9,434 | 8,735 | 699 | 472 | 227 |
| Stop lost | 351 | 249 | 102 | 89 | 13 |
| Start lost | 484 | 346 | 138 | 123 | 15 |
| In frame insertion | 1,774 | 1,557 | 217 | 130 | 87 |
| In frame deletion | 3,709 | 3,211 | 498 | 323 | 175 |
| Missense variant | 159,940 | 122,018 | 37,922 | 33,740 | 4,182 |
| Protein altering variant | 217 | 213 | 4 | 2 | 2 |
| Splice region variant | 14,766 | 10,966 | 3,800 | 3,441 | 359 |
| Incomplete terminal codon variant | 13 | 9 | 4 | 3 | 1 |
| Stop retained variant | 108 | 74 | 34 | 31 | 3 |
| Synonymous variant | 98,885 | 64,426 | 34,459 | 31,652 | 2,807 |
| Coding sequence variant | 24 | 18 | 6 | 5 | 1 |
| Mature miRNA variant | 84 | 52 | 32 | 28 | 4 |
| 5 prime UTR variant | 4,610 | 3,166 | 1,444 | 1,324 | 120 |
| 3 prime UTR variant | 9,188 | 6,242 | 2,946 | 2,648 | 298 |
| Non coding transcript exon variant | 17,351 | 11,545 | 5,806 | 5,151 | 655 |
| Intron variant | 54,488 | 37,776 | 16,712 | 15,124 | 1,588 |
| Upstream gene variant | 858 | 579 | 279 | 236 | 43 |
| Downstream gene variant | 410 | 263 | 147 | 133 | 14 |
| TF binding site variant | 5 | 4 | 1 | 1 | 0 |
| Regulatory region variant | 81 | 54 | 27 | 26 | 1 |
| Intergenic variant | 263 | 158 | 105 | 99 | 6 |

| **Supplementary Table S9.** Alternative allele frequencies (AAF) for common variants exclusively shared between Canary Islands and NAF populations, showing statistical differences in AAF both among islands and between the islands and the NAF population. | | | | | | | | | | | | |
| --- | --- | --- | --- | --- | --- | --- | --- | --- | --- | --- | --- | --- |
| **Chr** | **Position** | **Ref** | **Alt** | **rsID** | **EH** | **LP** | **LG** | **TF** | **GC** | **FV** | **LZ** | **NAF** |
| 6 | 117,113,762 | T | TA | rs371464745 | 0.129 | 0.032 | 0.125 | 0.076 | 0.097 | 0.109 | 0.111 | 0.116 |
| 7 | 6,713,940 | CAG | C | rs58381208 | 0.132 | 0.106 | 0.209 | 0.155 | 0.137 | 0.298 | 0.093 | 0.205 |
| 8 | 10,467,651 | C | CCTCTCTTCTT | rs773894295 | 0.033* | 0.035* | 0.072* | 0.070* | 0.072* | 0.064 | 0.071* | 0.214 |
| 8 | 23,282,562 | A | AGTATT | rs201841918 | 0.042 | 0.121 | 0.052 | 0.091 | 0.097 | 0.043 | 0.144 | 0.061 |
| 11 | 48,266,736 | C | G | rs7120775 | 0.208 | 0.106 | 0.049 | 0.097 | 0.158 | 0.032 | 0.086 | 0.091 |
| 13 | 100,517,195 | CTG | C | rs3831038 | 0.052 | 0.122 | 0.100 | 0.110 | 0.161 | 0.237 | 0.175 | 0.144 |
| 16 | 81,242,148 | GTT | G | rs55980345 | 0.491* | 0.379 | 0.321 | 0.355 | 0.381 | 0.426 | 0.355 | 0.259 |
| 17 | 39,595,484 | G | A | rs148768443 | 0.074 | 0.054 | 0.036* | 0.060 | 0.043* | 0.057 | 0.073 | 0.154 |
| 20 | 126,155 | GCAAA | G | rs11467497 | 0.325 | 0.242* | 0.184* | 0.226* | 0.223* | 0.234 | 0.232* | 0.438 |
| 20 | 126,310 | ACC | A | rs11467417 | 0.767 | 0.665 | 0.624 | 0.647 | 0.628 | 0.660 | 0.638 | 0.732 |
| 20 | 1,895,949 | A | AGT | rs148409797 | 0.193 | 0.332 | 0.322 | 0.278 | 0.306 | 0.319 | 0.407 | 0.307 |
| 20 | 1,895,950 | CCT | C | rs749337996 | 0.203 | 0.361 | 0.343 | 0.275 | 0.312 | 0.319 | 0.405 | 0.307 |
| 22 | 22,707,728 | C | T | rs148013584 | 0.125* | 0.066* | 0.173 | 0.146* | 0.111* | 0.202 | 0.147* | 0.320 |
| Alt, alternative allele; Chr, chromosome; EH, El Hierro; FV, Fuerteventura; GC, Gran Canaria; LG, La Gomera; LP, La Palma; LZ, Lanzarote; NAF, North African population; Ref, reference allele; TF, Tenerife. * Significant differences in AAF after Bonferroni correction with NAF population (*p* <0.001 for Fisher’s exact test). | | | | | | | | | | | | |

| **Supplementary Table S10.** Nonparametric Mann-Whitney U-test results for the principal components (PC) 1, PC2, and PC3 in PCA for Canary Islanders and the reference populations, excluding FIN and SSA. | | | | | | | | | | |
| --- | --- | --- | --- | --- | --- | --- | --- | --- | --- | --- |
| **PC1** | | | | | | | | | | |
|  | **CEU** | **GBR** | **IBS** | **EH** | **LP** | **LG** | **TF** | **GC** | **FV** | **LZ** |
| **GBR** | 0.001 | - | - | - | - | - | - | - | - | - |
| **IBS** | 5.35x10^-35^ | 9.70x10^-34^ | - | - | - | - | - | - | - | - |
| **EH** | 8.65x10^-35^ | 2.34x10^-33^ | 1.10x10^-33^ | - | - | - | - | - | - | - |
| **LP** | 8.44x10^-33^ | 1.58x10^-31^ | 2.31x10^-24^ | 1.68x10^-23^ | - | - | - | - | - | - |
| **LG** | 9.48x10^-39^ | 5.75x10^-37^ | 3.66x10^-40^ | 1.98x10^-33^ | 5.18x10^-36^ | - | - | - | - | - |
| **TF** | 9.78x10^-42^ | 1.15x10^-39^ | 1.05x10^-36^ | 9.91x10^-12^ | 2.37x10^-7^ | 2.12x10^-43^ | - | - | - | - |
| **GC** | 1.58x10^-45^ | 4.61x10^-43^ | 1.15x10^-44^ | 0.033 | 2.88x10^-25^ | 8.12x10^-29^ | 7.07x10^-16^ | - | - | - |
| **FV** | 3.99x10^-22^ | 1.47x10^-21^ | 5.48x10^-22^ | 1.63x10^-10^ | 8.05x10^-19^ | 1.02x10^-7^ | 4.54x10^-16^ | 3.04x10^-4^ | - | - |
| **LZ** | 1.79x10^-34^ | 4.56x10^-33^ | 1.64x10^-35^ | 2.21x10^-28^ | 6.42x10^-32^ | 4.54x10^-8^ | 1.48x10^-35^ | 6.92x10^-16^ | 0.006 | - |
| **NAF** | 6.91x10^-24^ | 3.18x10^-23^ | 1.75x10^-24^ | 2.88x10^-24^ | 2.61x10^-23^ | 7.67x10^-26^ | 2.42x10^-27^ | 8.14x10^-29^ | 1.96x10^-17^ | 4.39x10^-24^ |
| **PC2** | | | | | | | | | | |
| **GBR** | 0.005 | - | - | - | - | - | - | - | - | - |
| **IBS** | 4.49x10^-35^ | 8.86x10^-34^ | - | - | - | - | - | - | - | - |
| **EH** | 8.65x10^-35^ | 2.34x10^-33^ | 4.10x10^-36^ | - | - | - | - | - | - | - |
| **LP** | 1.12x10^-32^ | 1.58x10^-31^ | 3.50x10^-12^ | 1.78x10^-33^ | - | - | - | - | - | - |
| **LG** | 9.48x10^-39^ | 5.75x10^-37^ | 1.04x10^-37^ | 0.402 | 3.91x10^-32^ | - | - | - | - | - |
| **TF** | 9.78x10^-42^ | 1.15x10^-39^ | 8.03x10^-31^ | 9.69x10^-43^ | 7.22x10^-6^ | 2.31x10^-39^ | - | - | - | - |
| **GC** | 4.72x10^-45^ | 9.50x10^-43^ | 1.48x10^-12^ | 2.57x10^-44^ | 0.206 | 1.77x10^-45^ | 1.26x10^-11^ | - | - | - |
| **FV** | 3.99x10^-22^ | 1.47x10^-21^ | 1.61x10^-21^ | 6.62x10^-12^ | 2.88x10^-14^ | 2.27x10^-8^ | 6.35x10^-11^ | 1.87x10^-17^ | - | - |
| **LZ** | 1.79x10^-34^ | 4.56x10^-33^ | 9.58x10^-35^ | 9.32x10^-25^ | 5.35x10^-25^ | 3.28x10^-17^ | 2.43x10^-22^ | 4.62x10^-34^ | 0.773 | - |
| **NAF** | 3.32x10^-22^ | 1.36x10^-21^ | 9.30x10^-23^ | 6.50x10^-6^ | 1.13x10^-21^ | 0.001 | 2.35x10^-25^ | 5.37x10^-26^ | 3.05x10^-10^ | 1.23x10^-18^ |
| **PC3** | | | | | | | | | | |
| **GBR** | 0.979 | - | - | - | - | - | - | - | - | - |
| **IBS** | 1.06x10^-30^ | 4.29x10^-30^ | - | - | - | - | - | - | - | - |
| **EH** | 1.58x10^-33^ | 3.77x10^-32^ | 8.38x10^-35^ | - | - | - | - | - | - | - |
| **LP** | 5.18x10^-32^ | 4.95x10^-31^ | 1.65x10^-8^ | 2.50x10^-32^ | - | - | - | - | - | - |
| **LG** | 6.80x10^-14^ | 2.60x10^-13^ | 1.15x10^-14^ | 1.17x10^-17^ | 4.56x10^-14^ | - | - | - | - | - |
| **TF** | 2.47x10^-40^ | 1.63x10^-38^ | 9.84x10^-17^ | 2.05x10^-41^ | 0.068 | 1.36x10^-18^ | - | - | - | - |
| **GC** | 3.18x10^-42^ | 6.05x10^-40^ | 4.97x10^-44^ | 3.31x10^-45^ | 3.35x10^-37^ | 1.66x10^-35^ | 5.05x10^-50^ | - | - | - |
| **FV** | 3.96x10^-9^ | 6.46x10^-9^ | 2.05x10^-7^ | 3.78x10^-16^ | 2.03x10^-4^ | 1.77x10^-6^ | 4.53x10^-4^ | 4.46x10^-11^ | - | - |
| **LZ** | 7.71x10^-21^ | 5.17x10^-20^ | 2.21x10^-21^ | 3.23x10^-32^ | 2.10x10^-18^ | 2.14x10^-20^ | 4.30x10^-22^ | 0.207 | 1.14x10^-6^ | - |
| **NAF** | 3.32x10^-22^ | 1.36x10^-21^ | 9.30x10^-23^ | 1.48x10^-22^ | 1.13x10^-21^ | 4.63x10^-24^ | 2.41x10^-25^ | 2.12x10^-25^ | 3.31x10^-16^ | 1.30x10^-21^ |
| CEU, Utah Residents with Northern and Western European ancestry; EH, El Hierro; FV, Fuerteventura; GC, Gran Canaria; GBR, British; IBS, Iberian population in Spain; LG, La Gomera; LP, La Palma; LZ, Lanzarote; NAF, North African population; TF, Tenerife. Significant differences in PCs were established at *p* <9.09x10^-4^ considering a Bonferroni correction. | | | | | | | | | | |

| **Supplementary Table S11**. Genomic global ancestry proportions (%) obtained by ELAI in the Canary Islanders assuming 14 generations. | | | | | | | | | | | | |
| --- | --- | --- | --- | --- | --- | --- | --- | --- | --- | --- | --- | --- |
|  | | **EUR** | | | | |  | **NAF** |  |  | **SSA** |  |
| **Island** | | **Min** | | **Mean±SD** | | **Max** | **Min** | **Mean±SD** | **Max** | **Min** | **Mean±SD** | **Max** |
| El Hierro | | 69.9 | | 75.5±1.91 | | 81.7 | 16.9 | 22.2±1.76 | 27.6 | 1.29 | 2.34±0.55 | 4.04 |
| La Palma | | 77.0 | | 81.8±1.99 | | 86.9 | 12.2 | 16.0±1.61 | 19.9 | 0.90 | 2.26±0.81 | 4.72 |
| La Gomera | | 66.1 | | 73.7±2.07 | | 80.7 | 16.9 | 21.1±1.89 | 26.2 | 2.35 | 5.15±1.36 | 10.8 |
| Tenerife | | 74.5 | | 80.6±2.15 | | 85.2 | 12.3 | 16.9±1.83 | 21.2 | 0.77 | 2.48±0.77 | 5.59 |
| Gran Canaria | | 70.5 | | 79.4±2.39 | | 84.6 | 13.2 | 16.5±1.64 | 24.4 | 1.37 | 4.12±1.48 | 8.73 |
| Fuerteventura | | 69.9 | | 74.6±2.48 | | 79.8 | 17.4 | 21.8±1.91 | 25.9 | 1.95 | 3.62±0.97 | 6.05 |
| Lanzarote | | 69.6 | | 74.6±2.21 | | 81.6 | 16.0 | 21.5±1.85 | 26.0 | 2.06 | 3.95±0.87 | 6.28 |
| Total | | 66.1 | | 77.6±3.71 | | 86.9 | 12.2 | 18.9±3.10 | 27.6 | 0.77 | 3.52±1.51 | 10.8 |
| EUR, European; NAF, North African; SSA, sub-Saharan African. | | | | | | | | | | | | |
| **Supplementary Table S12.** Nonparametric Mann-Whitney U-test resulting p-values for the number of fragments in runs of homozygosity (ROHs) in the Canary Islands comparisons. | | | | | | | | | | | | |
|  |  | | **El Hierro** | | **La Palma** | **La Gomera** | | **Tenerife** | | **Gran Canaria** | **Fuerteventura** | |
|  | **La Palma** | | **0.002** | | - | - | | - | | - | - | |
|  | **La Gomera** | | **2.21x10^-5^** | | 0.228 | - | | - | | - | - | |
|  | **Tenerife** | | **5.89x10^-7^** | | 0.113 | 0.834 | | - | | - | - | |
|  | **Gran Canaria** | | **5.95x10^-12^** | | **2.15 x10^-4^** | 0.009 | | 0.010 | | - | - | |
|  | **Fuerteventura** | | **8.27x10^-7^** | | 0.003 | 0.030 | | 0.033 | | 0.551 | - | |
|  | **Lanzarote** | | **5.32x10^-15^** | | **1.37 x10^-7^** | **7.79x10^-6^** | | **4.11x10^-6^** | | 0.019 | 0.363 | |
|  | Significant differences after Bonferroni correction are shown in bold (established at *p*<2.38x10^-3^). | | | | | | | | | | | |

| **Supplementary Table S13.** Alternative allele frequency (AAF) per island for variants with AAF similar to the North African (NAF) population and different from the European (EUR) populations. | | | | | | |
| --- | --- | --- | --- | --- | --- | --- |
| **Chr** | 1 | 2 | 5 | 8 | 14 | 15 |
| **Position** | 151,006,539 | 135,911,422 | 33,963,870 | 19,816,934 | 102,349,907 | 28,510,895 |
| **Ref** | C | T | C | T | A | G |
| **Alt** | A | C | T | C | G | A |
| **rsID** | rs3738476 | rs17261772 | rs26722 | rs301 | rs2720207 | rs2240202 |
| **AAF** |  |  |  |  |  |  |
| **El Hierro** | 0.904 | 0.572 | 0.091 | 0.197 | 0.288 | 0.135 |
| **La Palma** | 0.864 | 0.495 | 0.103 | 0.239 | 0.304 | 0.185 |
| **La Gomera** | 0.883 | 0.635 | 0.120 | 0.222 | 0.301 | 0.135 |
| **Tenerife** | 0.851 | 0.540 | 0.168 | 0.264 | 0.276 | 0.143 |
| **Gran Canaria** | 0.880 | 0.512 | 0.082 | 0.274 | 0.363 | 0.137 |
| **Fuerteventura** | 0.924 | 0.554 | 0.130 | 0.152 | 0.326 | 0.130 |
| **Lanzarote** | 0.828 | 0.613 | 0.113 | 0.216 | 0.309 | 0.132 |
| AAF, alternative allele frequency; Alt, alternative allele; Chr, chromosome; Ref, reference allele. | | | | | | |

| **Supplementary Table S14**. Population Branch Statistic (PBS) for six prioritized variants with similar alternative allele frequencies (AAF) between Canary Islanders and the North African (NAF) population, but different from the European (EUR) populations. | | | |
| --- | --- | --- | --- |
| **Chromosome** | **Position** | **rsID** | **PBS** |
| 1 | 151,006,539 | rs3738476 | 0.303 |
| 2 | 135,911,422 | rs17261772 | 0.107 |
| 5 | 33,963,870 | rs26722 | 0.025 |
| 8 | 19,816,934 | rs301 | 0.133 |
| 14 | 102,349,907 | rs2720207 | 0.020 |
| 15 | 28,510,895 | rs2240202 | 0.003 |
| EUR includes GBR (British from England and Scotland), CEU (Utah Residents with Northern and Western European ancestry), and IBS (Iberian population in Spain) from 1KGP. | | | |

| **Supplementary Table S15**. Pairwise genetic distances (F_ST_) between Canary Islands, North African (NAF), and different European (EUR) populations in the six prioritized variants with similar alternative allele frequencies (AAF) between Canary Islanders and North African population, but differing from the European populations. | | | | | | | | | |
| --- | --- | --- | --- | --- | --- | --- | --- | --- | --- |
| **Variant** | | | **F_ST_ Canary Islands *vs.*** | | | | **F_ST_ NAF *vs.*** | | |
| **Chr** | **Position** | **rsID** | **CEU** | **GBR** | **IBS** | **NAF** | **CEU** | **GBR** | **IBS** |
| 1 | 151,006,539 | rs3738476 | 0.208 | 0.142 | 0.180 | 0.023 | 0.337 | 0.265 | 0.306 |
| 2 | 135,911,422 | rs17261772 | 0.214 | 0.169 | 0.063 | 0 | 0.206 | 0.161 | 0.057 |
| 5 | 33,963,870 | rs26722 | 0.096 | 0.072 | 0 | 0.009 | 0.166 | 0.141 | 0.033 |
| 8 | 19,816,934 | rs301 | 0.048 | 0.124 | 0.055 | 0.013 | 0.133 | 0.230 | 0.142 |
| 14 | 102,349,907 | rs2720207 | 0.015 | 0.105 | 0.047 | 0 | 0.015 | 0.105 | 0.046 |
| 15 | 28,510,895 | rs2240202 | 0 | 0.093 | 0.003 | 0.008 | 0.036 | 0.160 | 0 |
| CEU, Utah Residents with Northern and Western European ancestry (1KGP); Chr, chromosome; GBR, British from England and Scotland (1KGP); IBS, Iberian population in Spain (1KGP); NAF, North African population. | | | | | | | | | |

**Supplementary Table S16.** Functionally related genes in the region under putative positive selection and their associations with diseases and phenotypes. ^a^From Open Targets Platform; ^b^From the GWAS Catalog; ^c^The highest *p*-value was included. Available as a separate xls file.

**Supplementary Figures**

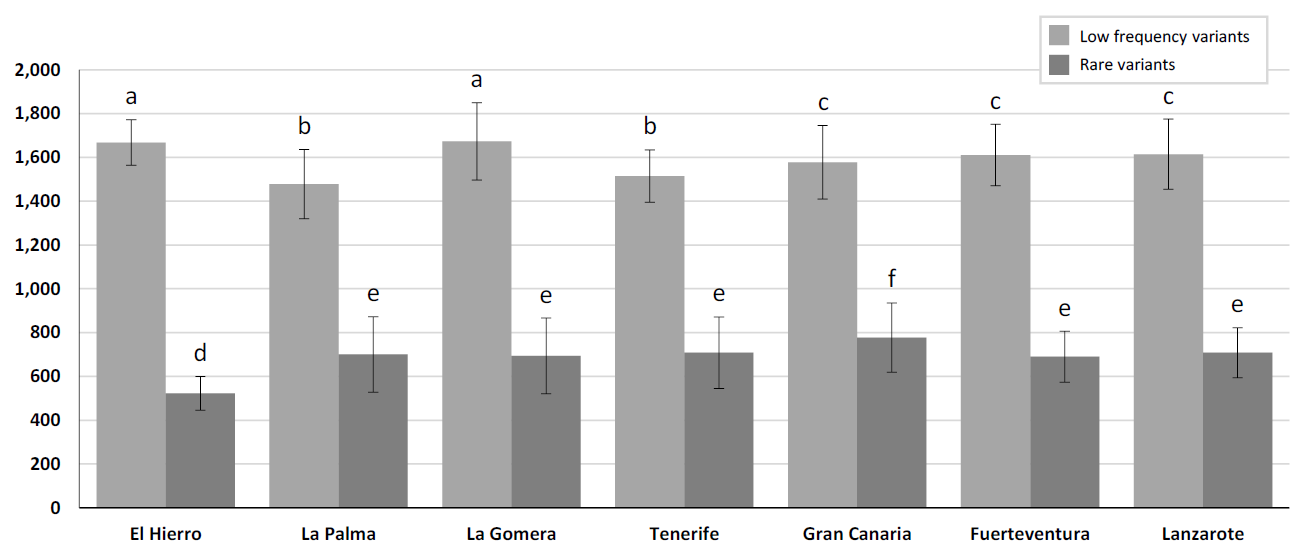

**Supplementary Figure S1**. Mean number of variants per island, per individual (± SD) within the two lowest allele frequency ranges. Islands with different letters indicate statistically significant differences in allele frequency, while the same letter indicates no significant differences between islands (Duncan's new multiple range test, p<0.05). Bars represent average ± SD.

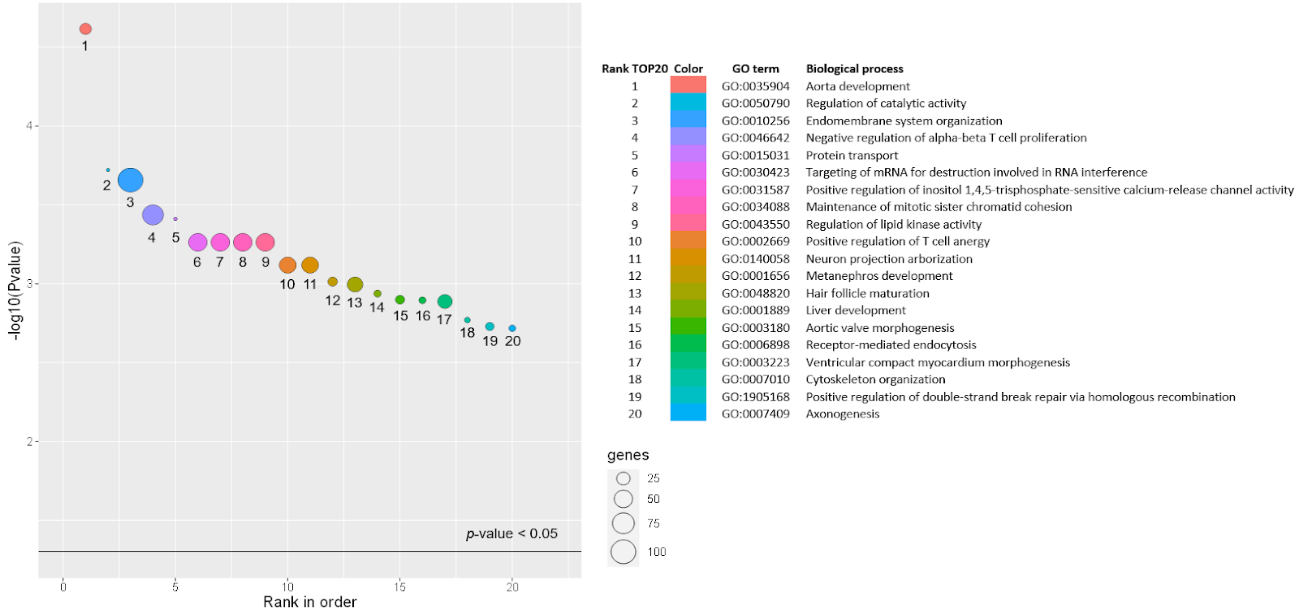

**Supplementary Figure S2**. Enrichment analysis of the 123 genes containing variants above the mean (> two variants) that meet the criteria for HC LoF, C-score >MSC and pLI >0.9. The size of the bubbles represents the percentage of functional genes covered by the user-defined gene list.

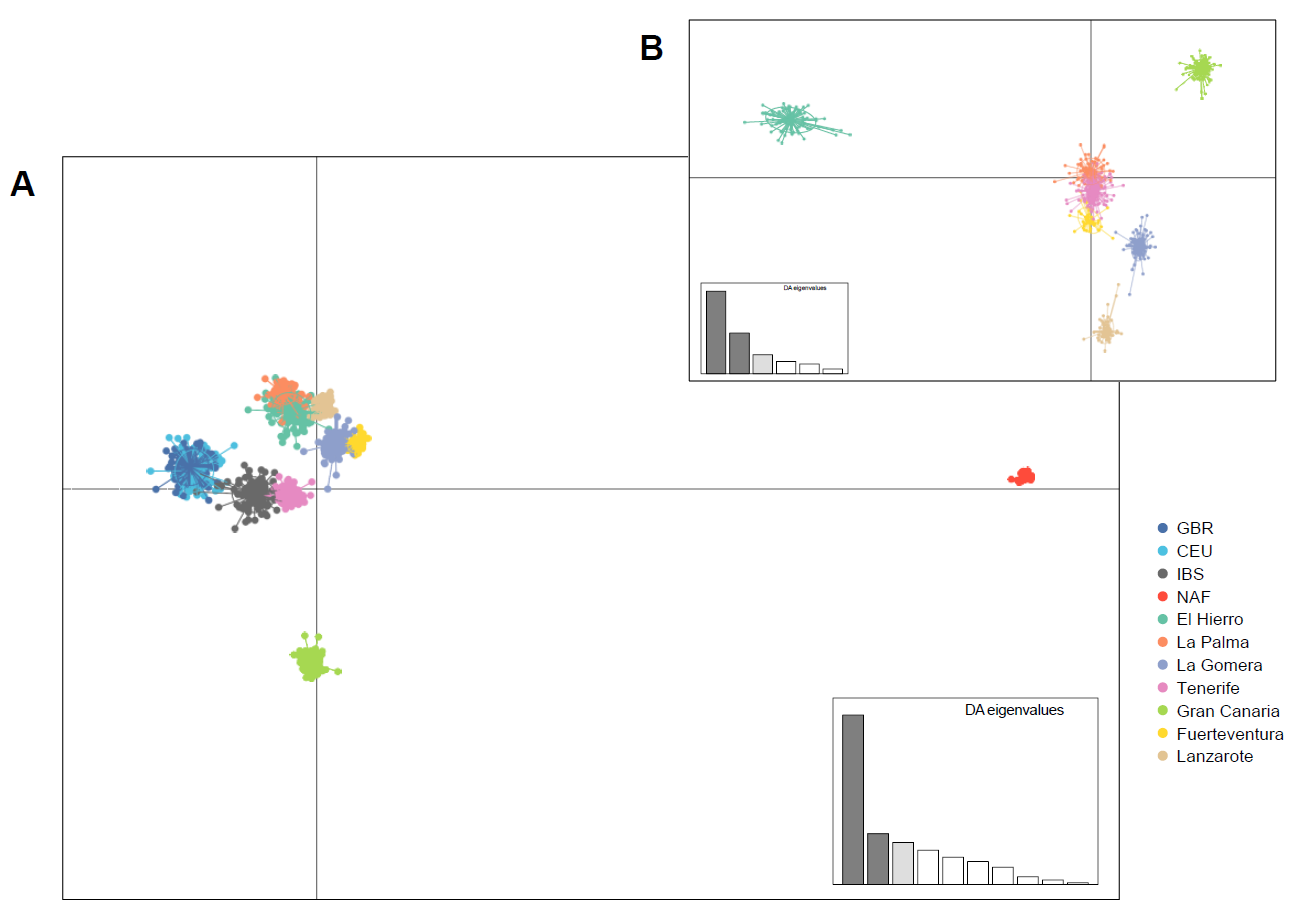

**Supplementary** **Figure S3.** Discriminant Analysis of Principal Components (DAPC) for rare variants (**A**) including Canary Islands and reference populations, excluding outlier populations (SSA and FIN); and (**B**) considering only Canary Islanders for rare variants. LD1, horizontal axis; LD2, vertical axis. CEU, Utah Residents with Northern and Western European ancestry (1KGP); GBR, British (1KGP); IBS, Iberian population in Spain (1KGP); NAF, North African population.

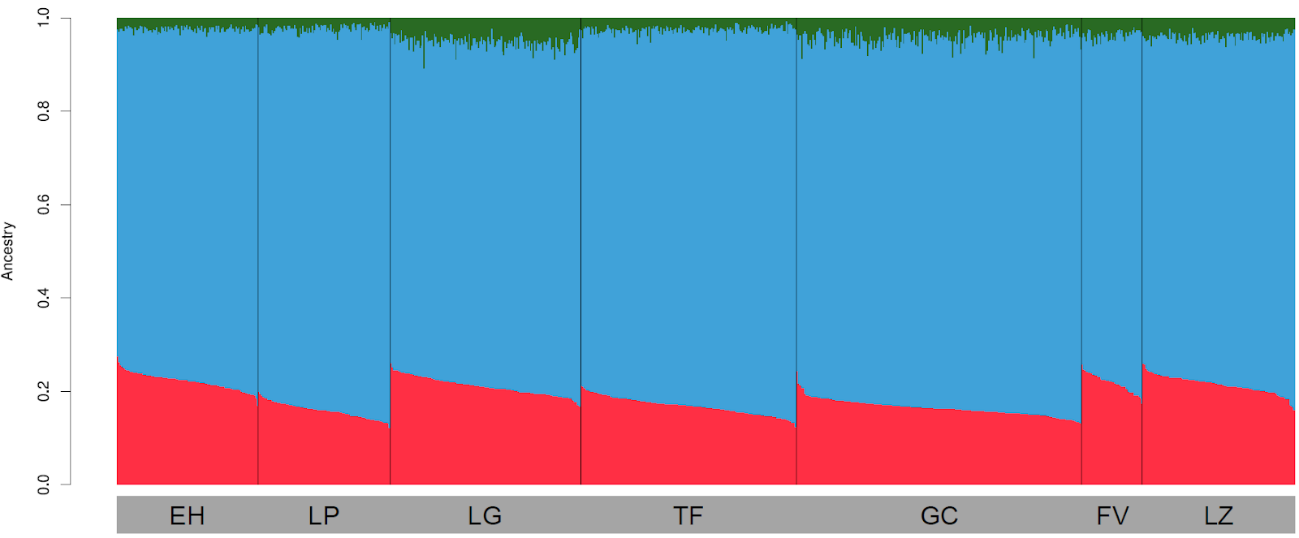

**Supplementary Figure S4.** Global ancestry estimates per island, per individual, based on a three-way admixture model of European (blue), North African (red), and sub-Saharan African (green) populations, as provided by ELAI. From left to right: El Hierro (EH), La Palma (LP), La Gomera (LG), Tenerife (TF), Gran Canaria (GC), Fuerteventura (FV), and Lanzarote (LZ).

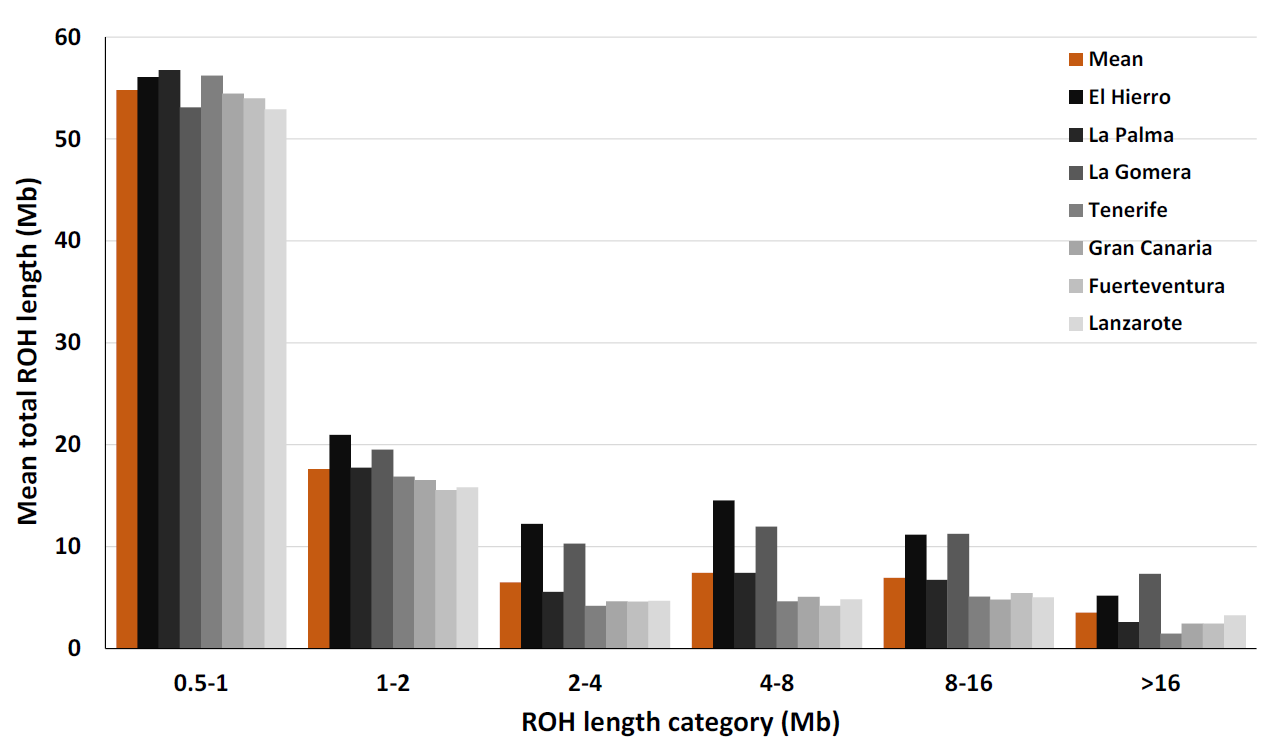

**Supplementary Figure S5**. Distribution of runs of homozygosity (ROHs) across ROH length classifications (0.5-1, 1-2, 2-4, 4-8, 8-16, and > 16 Mb) in the Canary Islands.

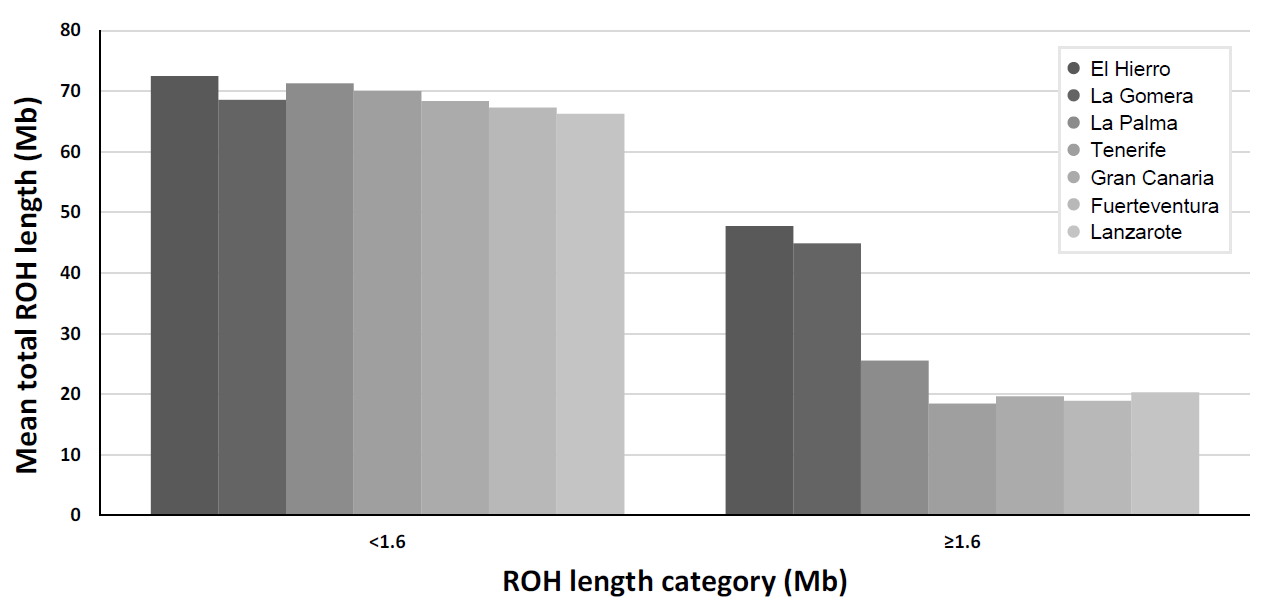

**Supplementary Figure S6**. Distribution of runs of homozygosity (ROHs) based on the modified Pemberton classification (<1.6 MB and ≥ 1.6 Mb) by average length (Mb) in the Canary Islands.
